# Interrogation of the Perturbed Gut Microbiota in Gouty Arthritis Patients Through *in silico* Metabolic Modeling

**DOI:** 10.1101/2020.09.02.20187013

**Authors:** Michael A. Henson

## Abstract

Recent studies have shown perturbed gut microbiota associated with gouty arthritis, a metabolic disease in which an imbalance between uric acid production and excretion leads to the deposition of uric acid crystals in joints. To mechanistically investigate altered microbiota metabolism in gout disease, 16S rRNA gene amplicon sequence data from stool samples of gout patients and healthy controls were computationally analyzed through bacterial community metabolic modeling. Patient-specific models were used to cluster samples according to their metabolic capabilities and to generate statistically significant partitioning of the samples into a *Bacteroides*-dominated, high gout cluster and a *Faecalibacterium*-elevated, low gout cluster. The high gout cluster samples were predicted to allow elevated synthesis of the amino acids D-alanine and L-alanine and byproducts of branched-chain amino acid catabolism, while the low gout cluster samples allowed higher production of butyrate, the sulfur-containing amino acids L-cysteine and L-methionine and the L-cysteine catabolic product H_2_S. The models predicted an important role for metabolite crossfeeding, including the exchange of acetate, D-lactate and succinate from *Bacteroides* to *Faecalibacterium* to allow higher butyrate production differences than would be expected based on taxa abundances in the two clusters. The surprising result that the high gout cluster could underproduce H_2_S despite having a higher abundance of H_2_S-synthesizing bacteria was rationalized by reduced L-cysteine production from *Faecalibacterium* in this cluster. Model predictions were not substantially altered by constraining uptake rates with different *in silico* diets, suggesting that sulfur-containing amino acid metabolism generally and H_2_S more specifically could be novel gout disease markers.

## Importance

Uric acid is produced in the human body from purine compounds contained in meat, poultry, and seafood. Gouty arthritic is a metabolic disease in which elevated levels of uric acid in the blood result in crystal formation, deposition in joints and chronic inflammation. Approximately 10 million people in the United States suffer from gout disease, and more than 2 million people take medications to lower blood uric acid levels. Recent experimental studies have shown that the human gut microbiota are perturbed in gout disease, suggesting that altered microbiota metabolism may result from gout development and/or treatment. Building on these experimental results, this study used computational metabolic modeling to investigate altered microbiota metabolism associated with gout disease. Patient-specific models were constructed and analyzed to predict microbiota-synthesized metabolites underproduced in gout patient versus healthy patients. The methodology identified butyrate, a well-known metabolite for promoting gut health, sulfur-containing amino acids and hydrogen sulfide, a metabolite known to promote inflammation, as possible metabolic markers of gout disease

## Introduction

The human gut microbiota play essential roles in digestion of plant polysaccharides (1, 2), synthesis of essential and health-promoting metabolites (3, 4), development of host immune response (5) and maintenance of colonization resistance to pathogens (6). The relative abundances of the diverse bacterial taxa that comprise the microbiota can be determined from stool samples through the application of 16S rRNA gene amplicon sequencing (7–9). As 16S sequencing has become increasingly routine, we have learned that numerous disease processes are correlated with disruptions of gut microbiota composition, also termed dysbiosis (10–12). Microbiota-associated diseases range from direct ailments of the gut such as inflammatory bowel disease (13) and *Clostridioides difficile* infection (14), to general metabolic diseases such as diabetes (15) and obesity (16), to systemic aliments such as cardiovascular disease (17), and even to neurological disorders such as depression (18) and Parkinson’s disease (19). While studies that correlate changes in microbiota abundances to disease development have revolutionized our understanding of human disease, such compositional-based analyses often provide little information about the underlying mechanisms by which the microbiota may drive and/or respond to disease processes.

Gouty arthritis is a metabolic disease related to the inability of the human host to properly regulate uric acid, a primary metabolite of purine metabolism (20–22). As the uric acid concentration in blood serum exceeds ∼400 μmol/L (termed hyperuricemia; (23, 24)), susceptible individuals may begin to suffer gout symptoms including painful inflammation due to the deposition of uric acid crystals in joints (25, 26). Therapeutic treatments include drugs such as Allopurinol and Febuxostat that reduce host uric acid synthesis, Krystexxa that increases the breakdown of uric acid to urea, Probenecid and Lesinurad which increase uric acid excretion, and a broad array of anti-inflammatory compounds (27, 28). Several recent studies in humans (29–31) and murine models (32–35) correlated changes in gut microbiota composition to the presence of gout disease, suggesting that microbiota properties could be used to monitor disease development, progression and recovery. Several of these 16S-based studies have been combined with gene catalog (36) and metabolomic (29) analyses to better understand the metabolic changes that accompanied compositional dysbiosis. While they provided new insights into the association between gout disease and altered gut microbiota, these studies were inherently limited in their ability to quantify the functional differences between the gut communities of gout patients versus healthy controls.

This *in silico* computational study was based on the hypothesis that altered gut microbiota were the result rather than the cause of gout disease, as a causative role has not been demonstrated to date. Indeed, uric acid in mainly produced in the liver by nucleic acid catabolism and only about 20% of uric acid production occurs from digestion of purine-rich foods (32–35). Furthermore, only about 30% of host generated uric acid is secreted into and excreted out of the intestine, with the remainder is excreted through the kidneys (35, 37). Although altered uric acid metabolism in the gut microbiota of gout patients has been demonstrated (36), such perturbations are unlikely to be the major cause of elevated uric acid levels in the blood. Consequently, this study examined the possibility of using predicted gut microbiota properties as clinically-relevant signatures of gout disease rather than as treatable disease drivers.

Consistent with this hypothesis, 16S abundance data derived from patient stool samples (36) were used to build sample-specific computational models for identifying microbiota-synthesized metabolites that may be under- or overproduced in gout patients compared to healthy controls. The 16S dataset, which included bacterial taxa abundances for 41 gout patients and 42 healthy controls, was processed using a metagenomics modeling pipeline (mgPipe; (38)) to construct community metabolic models that spanned 50 taxa (48 genera and 2 families). The computational models were simulated using three *in silico* diets, and the resulting simulation data was subjected to machine learning and statistical analyses to correlate metabolic function and patient type, extract information about metabolite synthesis capabilities at the community and individual taxa levels, predict intra-taxa metabolite crossfeeding relationships and explore the impact of dietary nutrient levels on community metabolism.

## Results

### Samples clustered by metabolic capability were associated with gouty and healthy patients

Prior to metabolic modeling, the 16S-derived abundance data were analyzed directly to identify community compositional features associated with gouty and healthy sample. Data analysis was limited to samples in which the modeled taxa accounted for at least 90% of the unnormalized abundances (39/41 gouty samples, 39/42 healthy samples; see Materials and Methods). Among the 25 most abundant taxa across the 78 sample, the abundances of six taxa were significantly different (Wilcoxon rank-sum test, FDR < 0.05) between the 39 gouty and 39 healthy samples. Notably, *Faecalibacterium* was significantly elevated (FDR = 3×10^−4^) in the healthy samples (average abundance 0.144) compared to the gouty samples (average abundance 0.063; Fig. S1A). Five taxa including three butyrate producers *(Faecalibacterium, Coprococcus, Roseburia)* were most negatively correlated with the blood uric acid concentration across the 78 samples (Fig. S1B). *Faecalibacterium* was most positively correlated to three butyrate producers *(Coprococcus, Roseburia, Subdoligranulum)* and *Akkermansia* as measured by the proportionality coefficient (Fig. S1C). A principal component plot of the taxa abundances showed no clear delineation of gouty versus healthy samples (Fig. S1D). Taken together, these results support the conclusion in the original experimental study (36) that depletion of *Faecalibacterium* and other butyrate producers was associated with gout development.

Using an *in silico* European diet (Table S2), the net maximal production rates (NMPCs) predicted for 409 exchanged metabolites across the 78 samples were clustered to investigate if model-predicted metabolic capabilities were associated with sample type. Clustering was performed within MATLAB using the kmeans method with the optimal number of three clusters (see Materials and Methods) and generated a silhouette value of 0.30 [doi:10.1016/j.ins.2015.06.039]. The three clusters produced a group of 26 samples dominated by *Bacteroides* (average abundance 0.75), a group of 44 samples with elevated *Faecalibacterium* (average abundance 0.15), and a small group of 8 samples with elevated *Prevotella* (average abundance 0.45; Fig. 1A). The *Bacteroides*-dominated cluster contained a disproportionately larger number of gouty samples (22/26) compared to the *Faecalibacterium*-elevated cluster (11/44, p < 10^−5^) and the entire sample set (39/78, p = 0.002; Fig. 1B). Similarly, the *Faecalibacterium*elevated cluster contained a disproportionate large number of healthy samples (33/44) compared to the entire sample set (39/78, p = 0.008). In terms of classification capability (doi: 10.9735/2229-3981), the *Bacteroides*-dominated cluster offered high precision with positive predictive value (PPV) = 0.85 but modest sensitivity with true positive rate (TPR) = 0.56. A principal component plot of the model-predicted NMPCs showed the *Prevotella*-elevated cluster samples as outliers and clearly identified the *Bacteroides*-dominated cluster samples as disproportionally gouty. Interestingly, gout patients have been reported to have elevated abundances of *Prevotella intermedia* in the oral microbiota (39). Taken together, these results suggested that elevated *Bacteroides* abundance may result from the gout disease process.

**Figure 1.**
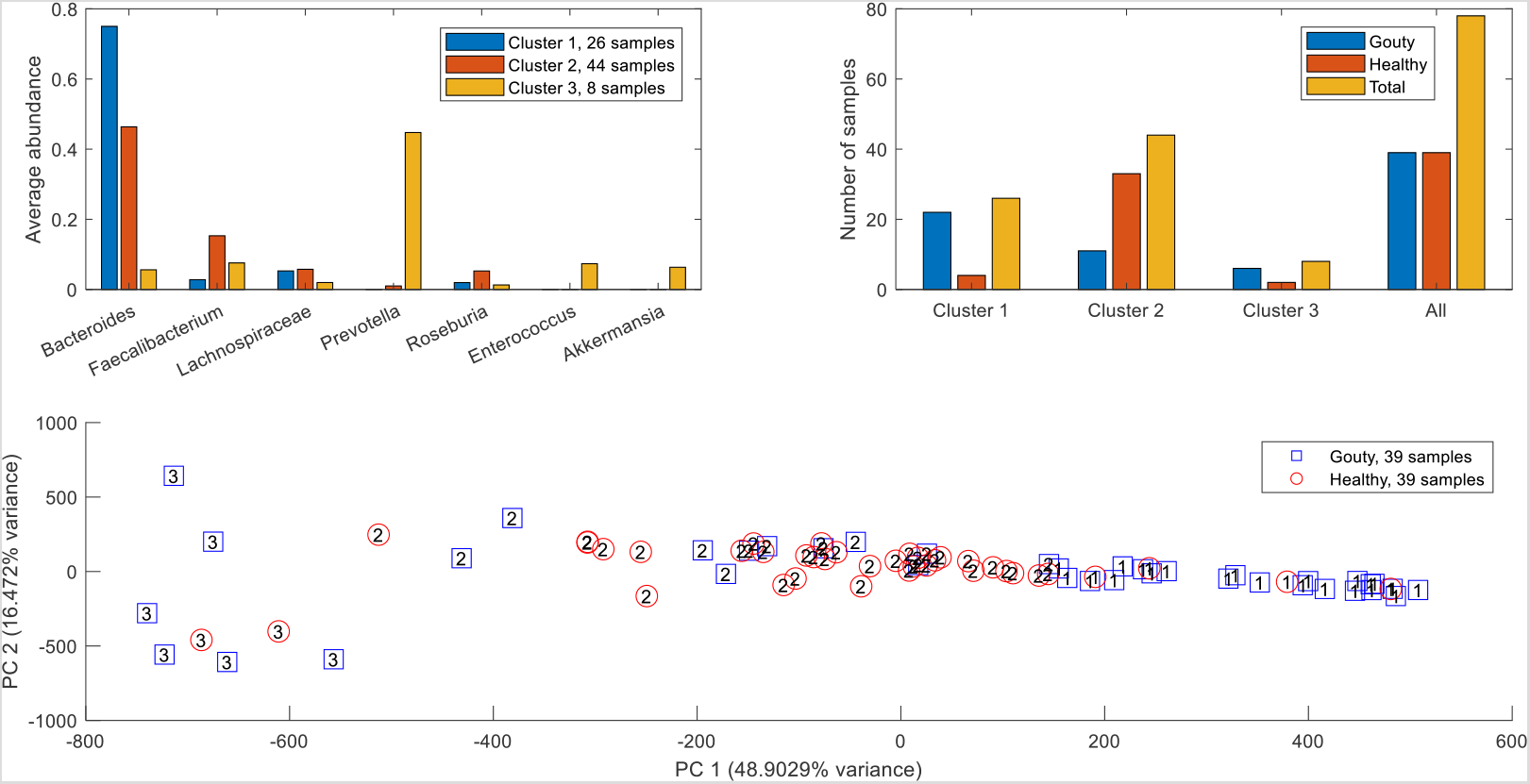
Clustering of model-processed 16S abundance data obtained with an average EU diet. (A) Average abundances of taxa which averaged at least 5% across at least one cluster. (B) Number of gouty, healthy and total samples in each cluster. Cluster 1 contained a disproportionate large number of gouty samples compared to cluster 2 (p < 10-5) and the entire dataset (p = 0.002). Cluster 2 contained a disproportionate large number of healthy samples compared to cluster 3 (p = 0.01) and the entire dataset (p = 0.008). (C) Principal component plot of the model-processed abundance data with gouty and healthy patient samples labeled by their associated clusters.

### Metabolic modeling predicted differential synthesis of amino acids and fermentation products in gouty and healthy samples

Due to the small number of samples contained in the *Prevotella*-elevated cluster, further statistical analyses were focused on the *Bacteroides*-dominated cluster (also called the high gout cluster) and the *Faecalibacterium*-elevated cluster (also called the low gout cluster). A rank sum test (FDR < 0.05), which assesses differences in median values, was performed to identify metabolites with the potential to be differentially produced in the high and low gout clusters. To reduce the 106 metabolites identified to a more manageable number, each metabolite also was required to have an average production rate of > 10 mmol/day in at least one cluster and to exhibit at least 10% difference between the average production rates in the two clusters. These thresholds ensured that the differential metabolites would have relatively high average production that differed between the two clusters. The resulting set of 42 differentially produced metabolites covered a wide range of metabolic pathways and included the amino acids D-alanine, L-alanine, L-cysteine, L-histidine, L-isoleucine, L-methionine and L-tyrosine as well as common products of gut microbiota fermentation such as butyrate, H_2_, H_2_S, isobutyrate, isocaproate, isovalerate and L-lactate (Figure S2, Table S6). Interestingly, hypoxanthine was the only the metabolite directly involved in purine metabolism that had the potential to be differentially produced between the two clusters, supporting the hypothesis that the gut microbiota were not the main drivers of gout disease. Similar predictions were obtained when the samples were partitioned directly according to their clinical status (Table S6), suggesting that sample clustering according to metabolite production capabilities captured the dominant metabolic features differentiating gouty and healthy samples.

Further computational analyses were performed on the seven differentially produced amino acids along with six additional amino acids that shared metabolic pathways with these seven amino acids and the eight differentially expressed fermentation products along with seven additional byproducts commonly produced by the gut microbiota. The high gout cluster was predicted to have significantly elevated capabilities for production of alanine, H_2_ and three products of branched-chain amino acid catabolism (isobutyrate from valine, isocaproate and isovalerate from leucine; Figure 2). By contrast, the low gout cluster was characterized by the potential for significantly elevated production of butyrate, L-lactate, the sulfur-containing amino acids L-cysteine and L-methionine, the L-cysteine catabolic product H_2_S, L-isoleucine and its catabolic product 3-methyl-2-oxovaleric acid, L-histidine and L-tyrosine. Model predictions of elevated alanine and reduced butyrate metabolism in gout patients compared to healthy controls were consistent with gene catalog and metabolomic studies (29, 31, 36).

**Figure 2.**
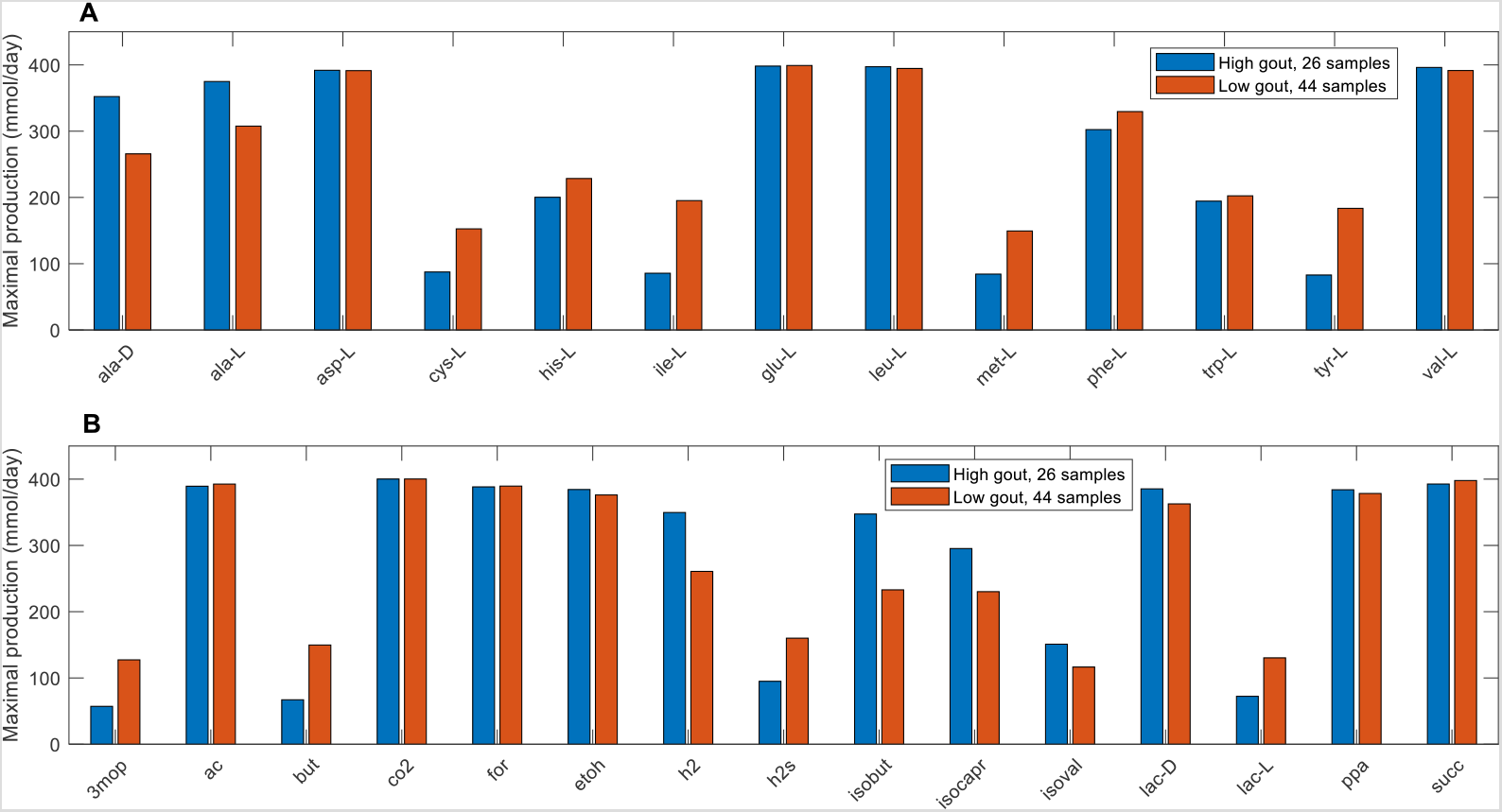
Maximal amino acid and fermentation byproduct synthesis capabilities in the high and low gout clusters from an average EU diet. (A) Classes of amino acids sharing common metabolic pathways and containing at least one amino acid differentially produced between the high and low gout clusters. (B) Common metabolic byproducts of carbohydrate fermentation and amino acid catabolism. Metabolite abbreviations are taken from the VMH database (www.vmh.life).

To further investigate the metabolite production capabilities of the gout- and health-associated gut communities, the contributions of individual taxa to the maximal synthesis of the differentially produced amino acids and fermentation byproducts were computed. *Bacteroides* was responsible for enhanced D-alanine, L-alanine and L-histidine production in the high gout cluster samples (Fig. 3), which were characterized by high *Bacteroides* abundances. The production of L-isoleucine and L-tyrosine, two amino acids not secreted by the *Bacteroides* metabolic model, were elevated in the low gout cluster due to increased synthesis by more abundant butyrate-producing taxa *(Faecalibacterium, Lachnospiraceae, Roseburia, Coprococcus)* as well as *Megamonas*. Interestingly, *Bacteroides* was predicted to have similar L-cysteine and reduced L-methionine synthesis capabilities in the high gout cluster despite these samples having relatively high *Bacteroides* abundances. This metabolic behavior resulted in significantly reduced total production of L-methionine and L-cysteine, which were also synthesized by *Faecalibacterium* and other butyrate producers, in the high gout cluster. These predictions suggested a possible role for sulfur-containing amino acids in the perturbed microbiota of gout patients. Synthesis of the six non-differentially produced amino acids was similar between the two clusters because elevated synthesis by *Bacteroides* in the high gout cluster was balanced with increased synthesis by butyrate producers and *Megamonas* in the low gout cluster (Fig. S3).

**Figure 3.**
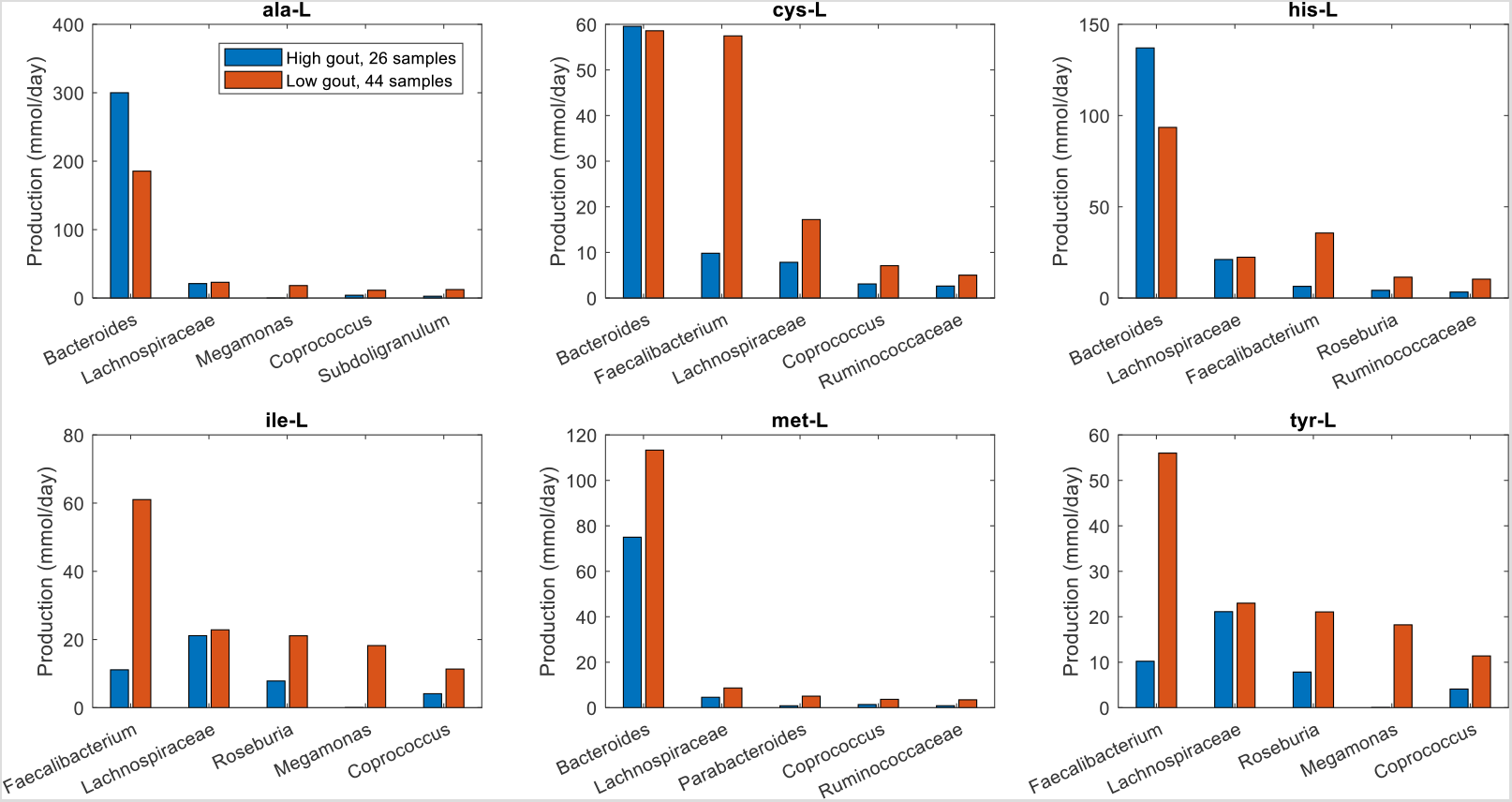
Individual taxa contributions to maximal synthesis of amino acids differentially produced between the high and low gout clusters from an average EU diet. The amino acids shown from top left to bottom right are L-alanine, L-cysteine, L-histidine, L-isoleucine, L-methionine and L-tyrosine. D-alanine has been omitted for brevity. For each amino acid, the top five taxa are shown in the order of their total production across the two clusters.

Similar analyses for differentially produced fermentation byproducts revealed that the potential for significantly elevated synthesis of H_2_, isobutyrate, isocaproate, and isovalerate in the high gout cluster was attributable to *Bacteroides* (Fig. 4). Byproducts not secreted by *Bacteroides* such as 3-methyl-2-oxovaleric acid, butyrate and L-lactate were predicted to have the potential for significantly elevated production in the low gout cluster due to increased synthesis by more abundant taxa. For example, butyrate was synthesized at higher rates by *Faecalibacterium, Roseburia, Coprococcus* and *Subdoligranulum* in the low gout cluster. In addition to the recognized importance of microbiota-derived butyrate for gut health (40, 41) and its previous implication as gout protective (36), these model predictions suggested that butyrate-producing taxa may contribute to the synthesis of other metabolites possibly involved in gut microbiota dysbiosis. For example, L-cysteine was predicted to have the potential for significantly elevated production in the low gout cluster due to enhanced synthesis by *Faecalibacterium* and other butyrate producers (Fig. 3). Since H_2_S is a common byproduct of cysteine degradation (42, 43), the ability of butyrate producers to synthesize L-cysteine could be related to the potential for elevated H_2_S production in the low gout cluster. These predictions along with previous studies showing H_2_S as a possible inducer of inflammation (44, 45) suggested a possible role for H_2_S specifically and sulfur-containing amino acids more generally in the perturbed microbiota of gout patients. As observed with amino acids, maximal production of the seven non-differentially produced byproducts was similar between the two clusters with elevated synthesis by *Bacteroides* in the high gout cluster being balanced with increased synthesis by butyrate producers and *Megamonas* in the low gout cluster (Fig. S4).

**Figure 4.**
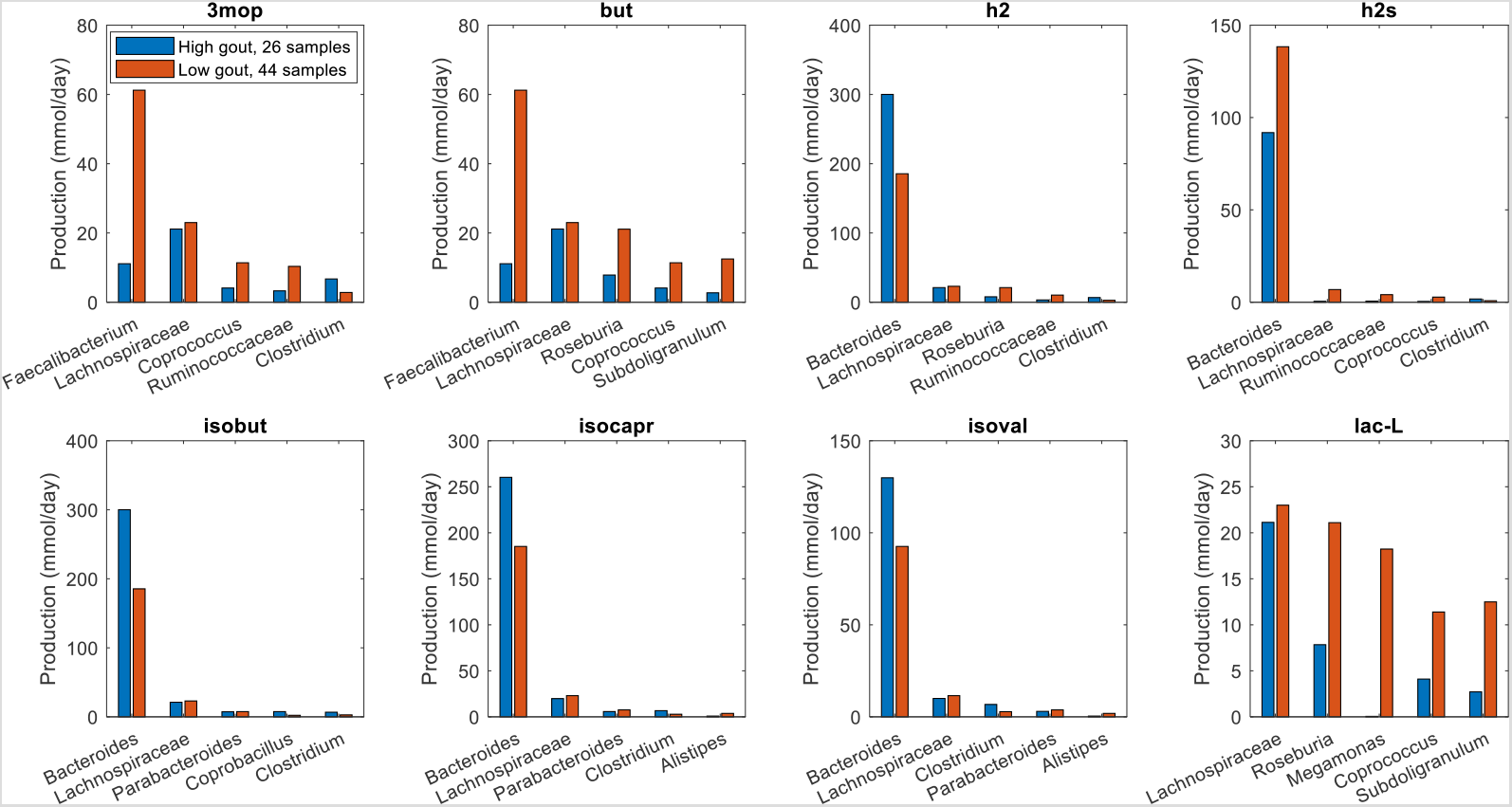
Individual taxa contributions to maximal synthesis of fermentation byproducts differentially produced between the high and low gout clusters from an average EU diet. The byproducts shown from top left to bottom right are 3-methyl-2-oxovaleric acid, butyrate, hydrogen, hydrogen sulfide, isobutyrate, isocaproate, isovalerate and L-lactate. For each byproduct, the top five taxa are shown in the order of their total production across the two clusters.

### Metabolite crossfeeding supports differential amino acid and fermentation byproduct synthesis in gouty and healthy samples

The previous model-based analyses identified butyrate and hydrogen sulfide as putative markers of gout disease. To investigate interactions between taxa that supported maximal production of these two metabolites, crossfeeding relationships were identified by finding metabolites which were both secreted by at least one taxa and uptaken by at least one other taxa above a defined threshold (5 mmol/day to focus on the largest contributors). Consistent with being more abundant in the low gout cluster, the five taxa mainly responsible for butyrate production were predicted to synthesize more butyrate in these sample communities (Fig. 4). However, butyrate production was higher than would be expected based on abundance differences between the two clusters. For example, *Faecalibacterium* was 230% more abundant in the low gout cluster (Fig. S1) yet synthesized 550% more butyrate. *Faecalibacterium* was predicted to achieve such elevated butyrate production by exploiting the availability of metabolites secreted from other taxa, most notably acetate, CO_2_, D-lactate and succinate from *Bacteroides* (Figure 5). Similarly, *Roseburia* utilized acetate and D-lactate from *Bacteroides* and *Subdoligranulum* utilized D-alanine from *Lachnospiraceae*. Very different crossfeeding relationships were predicted for maximal production of other fermentation byproducts. For example, *Bacteroides* exploited the availability of secreted L-alanine and format to achieve elevated D-lactate synthesis in the high gout cluster (Fig. S5). These results demonstrated the inherent metabolic flexibility of gut bacterial communities and suggested that taxa crossfeeding relationships could be highly context dependent.

**Figure 5.**
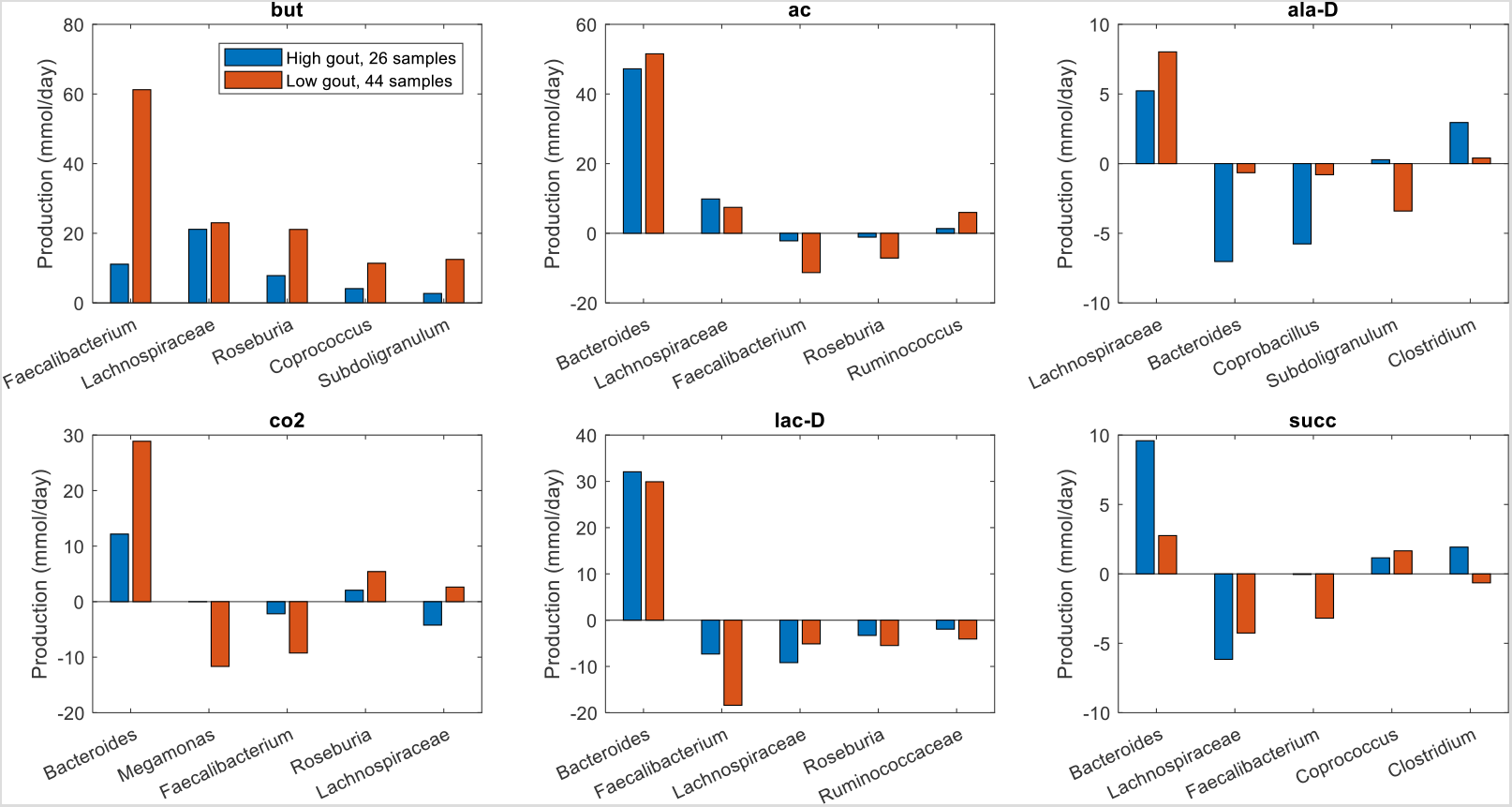
Individual taxa synthesis and uptake of crossfed metabolites for maximal butyrate production from an average EU diet. The metabolites shown from top left to bottom right are butyrate, acetate, D-alanine, carbon dioxide, D-lactate and succinate. Each metabolite shown had at least one taxa which satisfied a minimal bound on the metabolite secretion rate and the metabolite uptake rate. For each metabolite, the top five taxa were ordered by the sum of the absolute values of their uptake and secretion rates across the two clusters.

Unlike butyrate, H_2_S was predicted to be synthesized almost exclusively by a single taxa, *Bacteroides*. Although *Bacteroides* was 60% more abundant in the high gout cluster than the low gout cluster (Fig. 1), the maximal H_2_S production rate was 120% higher in the low gout cluster (Fig. 2). Because H_2_S is a product of cysteine degradation (46, 47) and maximal production of L-cysteine was elevated in the low gout cluster (Fig. 2), we hypothesized that L-cysteine crossfeeding was mainly responsible for differential H_2_S production between the two clusters. When H_2_S production was maximized, L-cysteine synthesis by *Faecalibacterium* was predicted to be 525% higher in the low gout cluster (Fig. 6), which matched the higher *Faecalibacterium* abundance in this cluster (Fig. 1). Elevated L-cysteine synthesis resulted in a 50% increase in H_2_S production by *Bacteroides*, which also preferentially utilized available D-lactate and L-lactate in the low gout cluster. Interestingly, D-alanine crossfeeding supporting maximal H_2_S production was predicted to differ dramatically between the clustered samples with *Bacteroides* consuming the metabolite in the high gout cluster and secreting the metabolite in the low gout cluster. *Faecalibacterium* was predicted to achieve maximal L-cysteine synthesis through acetate and D-lactate crossfeeding (Fig. S6). Collectively, these results demonstrated that complex relationships may exist between taxa and their metabolic products due to crossfeeding interactions that can be quantified with the type of metabolic modeling approach used in this study.

**Figure 6.**
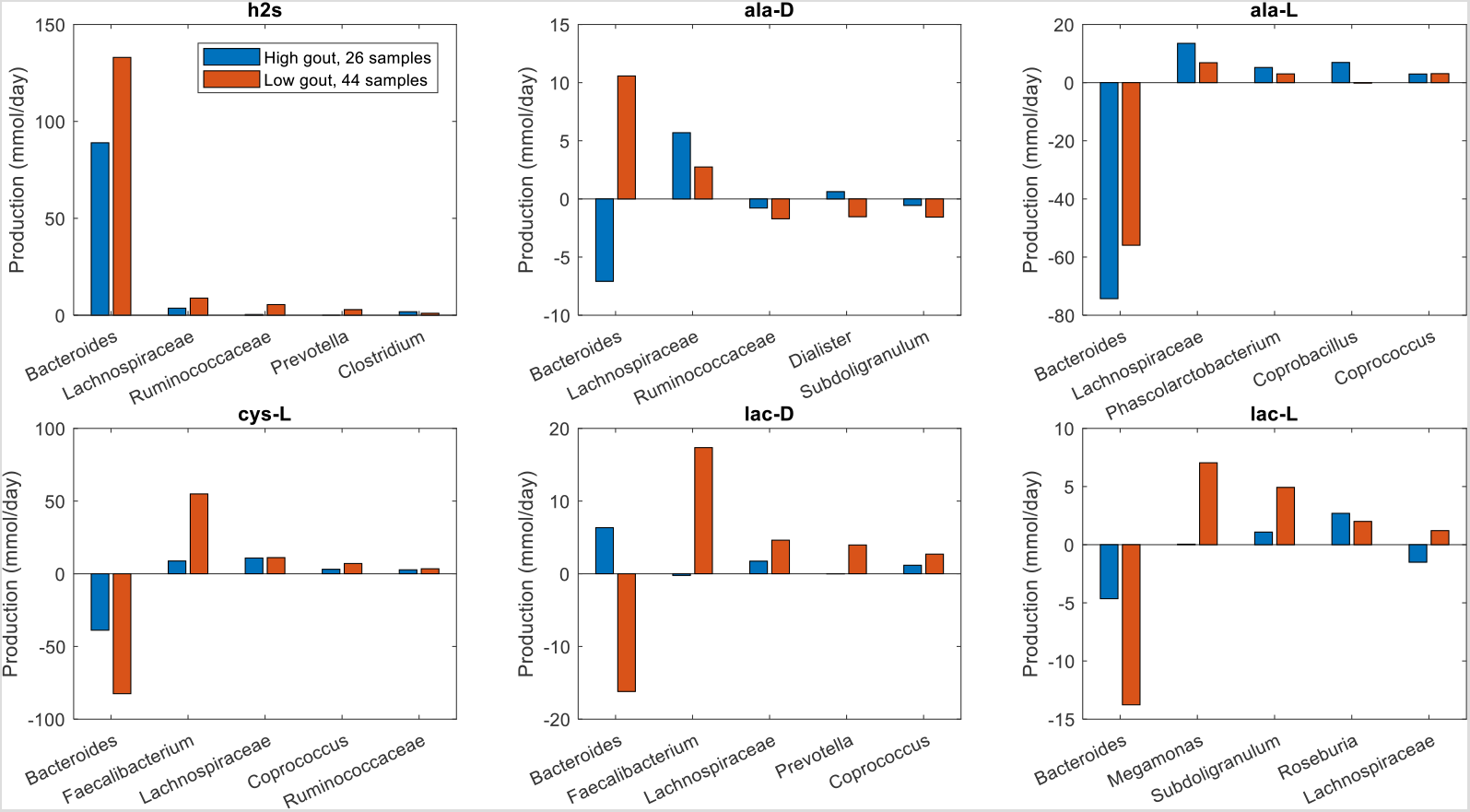
Individual taxa synthesis and uptake of crossfed metabolites for maximal H_2_S production from an average EU diet. The metabolites shown from top left to bottom right are hydrogen sulfide, D-alanine, L-alanine, L-cysteine, D-lactate and L-lactate. Each crossfed metabolite shown had at least one taxa which satisfied minimal bounds on the metabolite secretion and uptake rates. For each metabolite, the top five taxa were ordered by the sum of the absolute values of their uptake and secretion rates across the two clusters.

### Different in silico diets generated subtle changes in community metabolism

Previous simulations were performed by constraining community nutrient uptake rates according to an average EU diet. Diet is known to be strongly associated with gout disease, with high consumption of purine-rich foods such as meat, poultry and seafoods more likely to result in hyperuricemia and eventual gout development (48–50). To investigate the possible effects of dietary nutrients on microbiota metabolism, two other *in silico* diets were simulated (Table S2). Compared to the EU diet (EUD), the high protein diet (HPD) provided less total energy (1,792 vs. 2,372 kcal) with a lower proportion of energy coming from carbohydrates (37.4% vs. 41.2%) but a larger proportion coming from protein (28.7% vs. 11.9%) in the form of amino acids. By contrast, the high fiber diet (HFD) provided slightly more energy (2,436 kcal) with a larger contribution from carbohydrates (45.5%) and from protein (16.5%).

When model-processed abundance data generated with the HPD were clustered, three clusters (silhouette value 0.30) contained the same samples as when clustering was performed with the EU diet (Fig. S7A,B). However, when a rank sum test was performed to find metabolites with the potential to be differentially produced between the high gout clusters or the low gout clusters obtained with the two diets, 17 metabolites were identified including H_2_S and two metabolites associated with purine metabolism (guanosine, guanine; Fig. S7C). Otherwise, the two diets generated very similar clustered production of amino acids degradation and fermentation byproducts (Fig. S8). Collectively, these predictions suggested that *in silico* community metabolism was not strongly affected by changes in nutrient availability resulting from the simulated HPD.

Subtle changes in community metabolism were predicted when the EUD and HFD were compared. When partitioned with three clusters (silhouette value 0.29), the two diets generated different sample clustering with the high gout cluster increased from 26 to 31 samples and the low gout cluster decreased from 44 samples to 41 samples (Fig. 7A). The number of gouty samples in these two clusters also changed (Fig. 7B), but the *Bacteroides*-dominated cluster remained disproportionally gouty (23/31) compared to the *Faecalibacterium*-elevated cluster (12/41, p < 10^−3^) and the entire sample set (39/78, p = 0.03). As compared to EU diet, the *Bacteroides*-dominated cluster generated from the HFD had reduced precision with PPV = 0.74 but a slight increase sensitivity with TPR = 0.59. A rank sum test performed to find metabolites with the potential to be differentially produced between the high gout clusters or the low gout clusters of the two diets identified 10 metabolites, including three metabolites associated with plant polysaccharide degradation (D-galactose, D-glucose, D-maltose) and the glycolytic intermediate glycerol 3-phosphate (Fig. 7C). Interestingly, H_2_S was no longer differentially produced even though the HFD contained 84% more L-cysteine than the EUD as nutrient constraints. Although not statistically significant, the HFD produced slightly elevated maximal production of L-cysteine, L-histidine, L-isoleucine, 3-methyl-2-oxovaleric acid and L-lactate and slightly reduced maximal production of L-phenylalanine, H_2_, isobutyrate and isocaproate (Fig. S9). These predictions suggested subtle enhancements in sulfur-containing amino acid metabolism for the HFD and branched-chain amino acid catabolism for the EUD.

**Figure 7.**
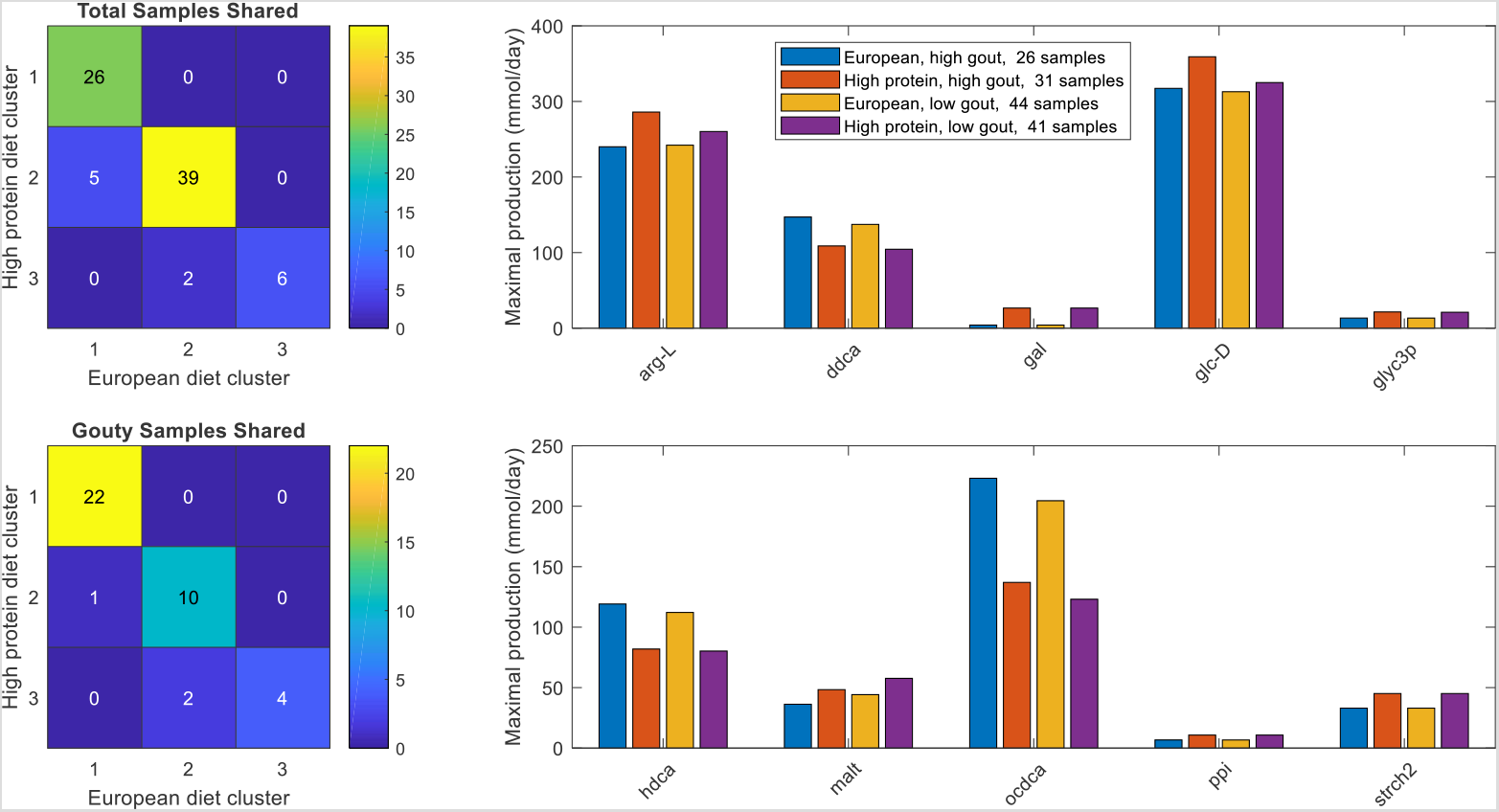
Sample clustering and differentially produced metabolites between average EU and high fiber diets. (A) Total samples shared between the three clusters obtained with the average EU diet and the three clusters obtained the high fiber diet. (B) Gouty samples shared between the three clusters obtained with the average EU diet and the three clusters obtained the high fiber diet. (C) Significant differences in maximal metabolite production rates were determined by applying the Wilcoxon rank sum test (FDR < 0.05) to each metabolite across all samples in the two high gout clusters and the two low gout clusters. In addition to being statistically different, each metabolite shown had an average production rate > 10 mmol/day in at least one of the compared clusters and average maximal production rates that differed between the compared clusters by at least 10%. Metabolite abbreviations are taken from the VMH database (www.vmh.life).

To further explore how dietary nutrients affected maximal H_2_S production, crossfeeding relationships were identified for the three high gout clusters and for the three low gout clusters generated from the different diets. When the low gout clusters were compared, the HPD was predicted to generate the most H_2_S due to elevated synthesis by *Bacteroides* (Fig. S10). Although the HPD contained the most dietary amino acids, the L-cysteine uptake rate by *Bacteroides* was not substantially different between the three diets. By contrast, the HPD generated only slightly higher H_2_S production than the HFD in the high gout clusters (Fig. 8). Interestingly, crossfeeding of L-cysteine from *Faecalibacterium* to *Bacteroides* was elevated for the HFD. These predictions suggested that H_2_S production was partially attributable to reactions associated with sulfur metabolism (51) other than L-cysteine degradation. The models predicted substantially reduced L-cysteine crossfeeding and H_2_S production in the high gout clusters compared to the low gout clusters across all three diets, reinforcing a possible role for cysteine catabolism and H_2_S production in the gout-perturbed microbiota.

**Figure 8.**
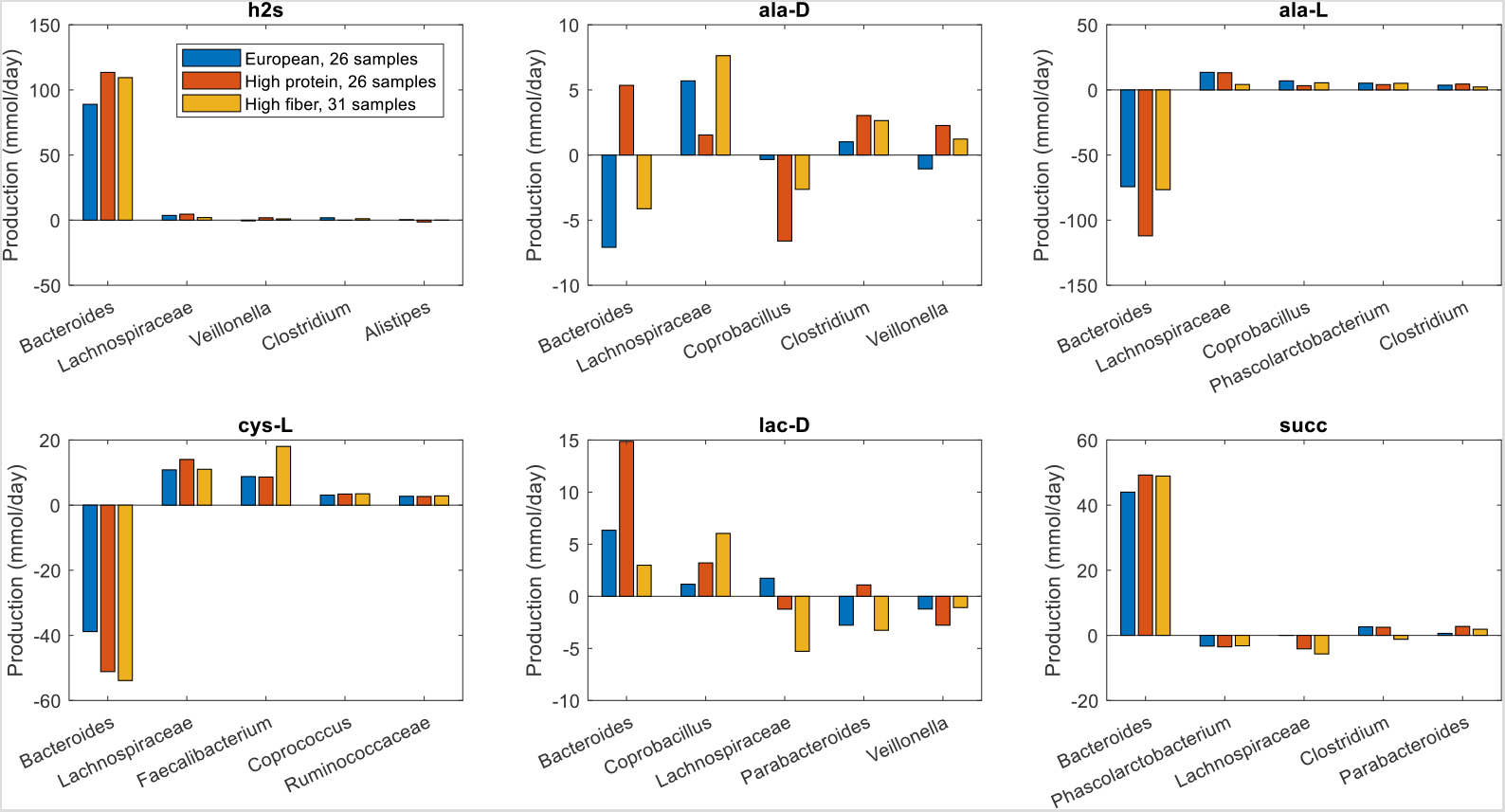
Individual taxa synthesis and uptake of crossfed metabolites for maximal H_2_S production in high gout clusters generated from average European, high protein and high fiber diets. The metabolites shown from top left to bottom right are hydrogen sulfide, D-alanine, L-alanine, L-cysteine, D-lactate and succinate. Each crossfed metabolite shown had at least one taxa which satisfied minimal bounds on the maximal metabolite secretion and uptake rates for at least one diet. For each metabolite, the top five taxa are ordered by the sum of the absolute values of their uptake and secretion rates across the three diets.

## Discussion

Gouty arthritis is a chronic inflammatory joint disease that results from imbalanced purine metabolism in the human host. Recent findings that the gut microbiota are perturbed by gout disease (30, 32–36) motivated this *in silico* modeling study aimed at identifying putative metabolic features associated with gout development. To this end, bacterial community metabolic models were constructed for 39 gout patients and 39 healthy controls using taxa abundance data generated from stool samples via 16S rRNA gene amplicon sequencing (36). Model simulations predicted the maximal possible production rates of 409 secreted metabolites for each of the 78 samples. By performing clustering analysis on these model-predicted metabolic capabilities, the samples were partitioned into a *Bacteroides*-dominated cluster with a disproportionate large number of gouty samples, a *Faecalibacterium*-elevated cluster with a disproportionate large number of healthy samples, and a *Prevotella*-dominated cluster with only six samples. Consistent with the original experimental study (36), these predictions suggested that elevated *Bacteroides* and reduced *Faecalibacterium* abundances were signatures of gout disease.

To gain mechanistic insights into gut metabolic features associated with gout, secreted metabolites with significantly different maximal synthesis rates in the *Bacteroides*-dominated, high gout cluster and the *Faecalibacterium*-elevated, low gout cluster were identified for a simulated EU diet. Interestingly, hypoxanthine was the only identified metabolite associated directly with purine metabolism. Of the four purine bases (adenine, guanine, hypoxanthine, xanthine), foods rich in hypoxanthine such as animal and fish meats have been reported to be more strongly associated with gout development (50). This uric acid precursor was elevated in the low gout cluster, suggesting higher rather than lower synthesis capabilities for uric acid, which was not secreted by the metabolic models. By contrast, the low gout cluster was predicted to have elevated synthesis of many metabolites, most notably gut health promoting butyrate and several metabolites associated with sulfur-containing amino acid metabolism including the L-cysteine degradation product H_2_S. Although altered uric acid metabolism in the gut microbiota of gout patients has been demonstrated (36), the *in silico* model predictions suggested that such perturbations are unlikely to be the major cause of elevated uric acid levels in the blood and that altered gut microbiota were the result rather than the cause of gout disease.

Detailed analyses of individual taxa contributions to the maximal synthesis of differentially produced amino acids predicted a tradeoff between *Bacteroides* and butyrate producers such as *Faecalibacterium, Roseburia, Subdoligranulum* and *Coprococcus*. D-Alanine and L-alanine elevated in the high gout cluster were synthesized primarily by *Bacteroides*, while amino acids elevated in the low gout cluster were not secreted by *Bacteroides* (L-isoleucine, L-tyrosine) or synthesis was more dependent on butyrate producers (L-cysteine, L-histidine, L-methionine). Enhanced alanine metabolism in gut communities of gout patients has been proposed previously based on gene catalog (36) and metabolomics (29) analyses. Model predictions associated with L-cysteine and L-methionine were novel and particularly interesting since *Bacteroides* was more abundant in the high gout cluster yet was predicted to have lower maximal synthesis rates of these two sulfur-containing amino acids.

Similar analyses performed for maximal synthesis of common fermentation byproducts predicted elevated synthesis of isobutyrate, isocarpoate and isovalerate in the high gout cluster resulting from branched-chain amino acid catabolism. The low gout cluster was predicted to have elevated maximal production of the short-chain fatty acid butyrate, L-lactate and H_2_S, a common end product of L-cysteine catabolism. The ability of bacterial communities contained in the low gout cluster to generate higher butyrate levels was related to higher abundances of butyrate producers in these samples as well as the crossfeeding of metabolites such as acetate, D-alanine, D-lactate and succinate to the butyrate producers. Butyrate has been widely identified as a gut health promoting metabolite (40, 41), and its reduced production by the gut microbiota has been associated with gout disease in several other studies (31, 36). Unlike some previous studies, the *in silico* models did not predict that propionate (31, 36) would be reduced or that acetate would be elevated (29) in the gut communities of gout patients. Therefore, the computational predictions support the hypothesis that butyrate is the key short-chain fatty acid associated with gout development and that loss of butyrate producers may an important feature of gout-altered gut microbiota.

Reduced maximal H_2_S production by *Bacteroides* in the high gout cluster was consistent with the prediction of lower total L-cysteine synthesis in this cluster. By contrast, the low gout cluster had elevated maximal H_2_S production due to substantially increased L-cysteine crossfeeding from *Faecalibacterium* to *Bacteroides*. While *Bacteroides* is not typically viewed as an important genus for H_2_S production (46), many *Bacteroides* strains process the necessary enzymes for cysteine-to-H_2_S conversion (52, 53). H_2_S has been proposed to have both anti-inflammatory and proinflammatory effects on the human host depending on a number of increasingly understood factors (54–56), including whether H_2_S is synthesized endogenously by mucosal epithelial cells or derived from the gut microbiota (43, 57). More specifically, H_2_S synthesized by the gut microbiota through cysteine degradation has been proposed to promote gut health at relatively low concentrations (47). Since most cysteine-to-H_2_S conversion in the gut is performed by the microbiota (47), these results suggested that tight regulation of this metabolic process may be necessary to avoid deleterious effects associated with elevated H_2_S concentrations. More generally, the *in silico* predictions revealed an interesting gut community behavior where synthesis of a potentially inflammatory metabolite (e.g. H_2_S) may be supported by a health-promoting taxa (e.g. *Faecalibacterium)* crossfeeding a metabolite (e.g. L-cysteine) to a disease-promoting taxa (e.g. *Bacteroides)*. Collectively, these results suggest that cysteine-to-H_2_S conversion by the gut microbiota may represent a novel metabolic signature of gout disease.

Because consumption of high-purine containing foods such as meat, poultry and fish is a known correlative to gout disease (48–50), *in silico* high protein and high fiber diets were simulated and compared to the average EU diet. The high protein diet (HPD) allowed higher amino acid and lower carbohydrate uptake rates and therefore was expected to generate substantially different predictions than the EU diet (EUD). This hypothesis proved to be largely false, although H_2_S continued to be elevated in the low gut cluster with the HPD. Similar results were obtained when the high fiber diet (HFD) was compared to the EUD, with the exception that sample clustering was altered between the two diets. These inconclusive results may have been attributable to limitations of the *in silico* approach in which sample community compositions were fixed while dietary nutrients were varied, while in reality different diets would be expected to alter microbiota composition. A more consistent analysis could be performed by having 16S-derived abundance data for both gout patients and healthy controls over a range of known diets. Despite this limitation, one consistent prediction across all three diets was elevated H_2_S in the low gout clusters. Therefore, these results again supported enhanced cysteine-to-H_2_S conversion as a novel metabolic signature of gout disease.

## Materials and Methods

### Patient Data

Gut microbiota composition data were obtained from a published study (36) in which stool samples from 83 patients were subjected to 16S rRNA gene amplicon library sequencing. The study included 41 gout patients as determined by clinical symptoms and elevated blood uric acid levels and 42 healthy controls. The patients ranged in age from 27 to 75 years (average 49.1 years) and contained 41% females. For each patient sample, 16S-derived relative bacterial abundances were provided at different taxonomic levels that included 15 phyla, 28 classes, 38 orders, 71 families and 129 genera. Extensive clinical metadata, including the blood uric acid concentration, also were provided for each sample (Table S1).

### Community Metabolic Modeling

Community metabolic models were restricted to 50 taxonomic groups to limit the computational effort associated with model building, simulation and analysis while ensuring adequate coverage of the 16S OTU read data. The models accounted for the 48 most abundant genera across the 83 samples subject to the requirement that each genus could be modeled using genome-scale metabolic reconstructions available in the Virtual Metabolic Human (VMH) resource ((58); www.vmh.life). Combined reads for *Escherichia*/*Shigella* were equally split between the two genera. Because unidentified *Lachnospiraceae* and *Ruminococcaceae* accounted for 4.9% and 1.1% of total reads, respectively, these two families were included in the models and combined with unmodelable genera *(Lachnobacterium, Anaerosporobacte*r, *Parasporobacterium, Hespellia* and *Robinsoniella* for *Lachnospiraceae*; *Oscillibacter, Anaerofilum, Acetivibrio, Acetanaerobacterium, Sporobacter* and *Hydrogenoanaerobacterium* for *Ruminococcaceae)*. This procedure resulted in 50 modeled taxa that accounted for an average of 97.0% of total OTU reads across the 83 samples (Table S1).

For each sample, the OTUs associated with each taxon were summed and then normalized to unity by dividing by the sum of OTUs. These normalized taxa abundances were used to construct sample-specific community metabolic models. The function createPanModels within the metagenomics pipeline (mgPipe; (38)) of the MATLAB Constraint-Based Reconstruction and Analysis (COBRA) Toolbox (59) was used to construct genus- and family-level models from the 818 strain models available in the VMH database. According to function documentation available on the COBRA website, the function createPanModels combined all reactions for strains belonging to the taxon of interest and attempted to remove futile cycles that may result from the combined reactions by making certain reactions irreversible. Because the 16S data did not provide resolution at the species and strain levels, the community metabolic models were unable to account for differences in community composition and function below the genus level. Despite this limitation, the results showed that the pan-genome metabolic models used for community modeling allowed substantial differentiation of samples according to their functional capabilities.

The function initMgPipe was used to construct a community metabolic model for each of the 83 patient samples. Model construction required specification of taxa abundances for each sample and maximum uptake rates of dietary nutrients, which were specified according to EU average, high protein and high fiber diets downloaded from the VMH resource (Table S2). The community models contained an average of 38,570 reactions (minimum 22,099; maximum 57,820). All models contained the same constraints for the maximum nutrient uptake rates specified for the chosen diet, while each model had different constraints imposed for the sample taxa abundances.

Following the model building process, mgPipe automatically performed flux variability analysis (FVA) for each model with respect to each of the 409 metabolites assumed to be exchanged between the microbiota and the lumen and fecal compartments. FVA calculations were performed with the COBRA code fastFVA using the CPLEX linear program solver to either maximize/minimize the production of the metabolite or to maximize/minimize the uptake of the metabolite subject to the additional constraint that the community biomass flux remained in the range 0.4-1.0 mmol/day (60). The FVA results were used to compute the net maximal production capability (NMPC; (60)) of each metabolite by each model. Each NMPC value was calculated as the difference between the objective functions of two computed FVA solutions, the first which maximized metabolite production/secretion into the fecal compartment and the second which minimized metabolite uptake from the lumen compartment. An NMPC value represents the sample-specific potential for community production of a single metabolite given the applied nutrient uptake and biomass flux constraints. The mgPipe framework (38) is based on analysis of metabolite-specific NMPCs across samples to assess the capabilities of modeled communities to differentially produce metabolites. Due to nature of the FVA calculations, the NMPCs do not imply that maximal production of multiple metabolites may be achieved simultaneously. In this study, each NMPC was calculated and analyzed independently for each modeled sample. Furthermore, NMPCs were calculated for the three different diet to assess the possible impact of nutrient levels on community metabolism (Tables S3-S5).

Unfortunately, mgPipe did not offer the capability to directly extract the metabolite synthesis capabilities of each modeled taxa. This information was important to understand which taxa were contributing to metabolite synthesis and the intra-taxa crossfeeding relationships which supported maximal production of a particular metabolite. Therefore, specialized MATLAB scripts were written to utilize available Microbiome Modeling Toolbox functions (e.g. adaptVMHDietToAGORA, guidedSim, useDiet) to perform FVA with respect to selected metabolites and to extract taxa-level secretion and uptake fluxes for relevant metabolites. The complete metabolic modeling workflow can be viewed as complementary to more established metagenomic analysis techniques such as Phylogenetic Investigation of Communities by Reconstruction of Unobserved States (PICRUSt; (61)) by quantifying community interactions such as nutrient competition, metabolite crossfeeding and product synthesis.

### Data analysis

Patient data consisted of normalized taxa abundances and model-predicted data consisted of diet-dependent NMPCs, both of which could be connected to associated metadata on a sample-by-sample basis (Table S1). Data analysis was limited to samples in which the modeled taxa accounted for at least 90% of the unnormalized abundances (39/41 gouty samples, 39/42 healthy samples) to achieve adequate representation of the original 16S gene amplicon data. Both normalized taxa abundances and model-predicted NMPCs were subjected to unsupervised machine learning techniques including clustering and principal component analysis (PCA) to extract putative relationships between partitioned samples and patient gout status. Rather than apply supervised learning to samples partitioned on their known clinical status (i.e. gouty, healthy), unsupervised learning was performed to determine if samples clustered by taxa abundances or NMPCs could be associated with gout status. This approach was applied under the hypothesis that clustering could partially unravel the complex gout disease etiology and reveal at least one cluster with statistically high levels of gouty or healthy samples.

Clustering was performed using the MATLAB function kmeans with the squared Euclidean distance metric, the k-means++ algorithm for cluster center initialization (62) and 1,000 replicates. When clustering was applied to NMPC data generated from the average EU diet, an optimal number of three clusters was determined using the MATLAB function evalclusters with the kmeans clustering method, the silhouette evaluation criterion (63), the sum of absolute difference as the distance measure and 100 replicates. To facilitate subsequent comparisons, three clusters also were used for the high protein and high fiber diets and for clustering of abundance data. For each *in silico* diet tested, the clustering method proved robust in that the same clustered samples were consistently returned despite the randomness of cluster initialization and the existence of local minima (64).

PCA was performed directly on normalized taxa abundances and model-predicted NMPC data rather than on data preprocessed with sample dissimilarity measures such as the Bray–Curtis (65) or UniFrac (66) metrics. This approach was deemed appropriate since PCA was used for preliminary data visualization and not quantitative data analysis. Statistical significance of associations between categorial variables (e.g. gouty/healthy) across sample groups were assessed using Fisher’s exact test (67). Correlations between taxa based on their abundances across samples were calculated using the proportionality coefficient (68), which accounts for the effects of data normalization. Statistically significant differences between NMPCs across samples were assessed using the Wilcoxon rank-sum test (69). The resulting p-values were used to calculate the false-positive discovery rate (FDR) for each metabolite using the MATLAB function mafdr with the Benjamini-Hochberg method (70).

## Data Availability

All data used in model development and analysis are provided in the Supplemental Materials. All MATLAB codes will be provided as the Supplemental Materials if the manuscript is accepted for publication.

## Acknowledgements

This research received no specific grant from any funding agency in the public, commercial or not-for-profit sectors. The author wishes to acknowledge his Ph.D. student Ayushi Patel for her assistance with generating the references.

---

**Table 1.** Summary of patient metadata from (36).

|  | **Gouty** | **Healthy** | **Total** |
| --- | --- | --- | --- |
| Patients | 41 | 42 | 83 |
| Age (years) | 49.4 | 48.7 | 49.1 |
| Female/Male | 17/24 | 17/25 | 34/49 |
| Body Mass Index<br>(kg/m<sup>2</sup>) | 23.1 | 23.2 | 23.2 |
| Blood Uric Acid<br>(μmol/L) | 496 | 242 | 368 |
| Urea Nitrogen<br>(mmol/L) | 7.78 | 4.48 | 6.11 |
| Blood Glucose<br>(mmol/L) | 5.76 | 5.17 | 5.46 |

## Supplementary Information

**Figure S1.**
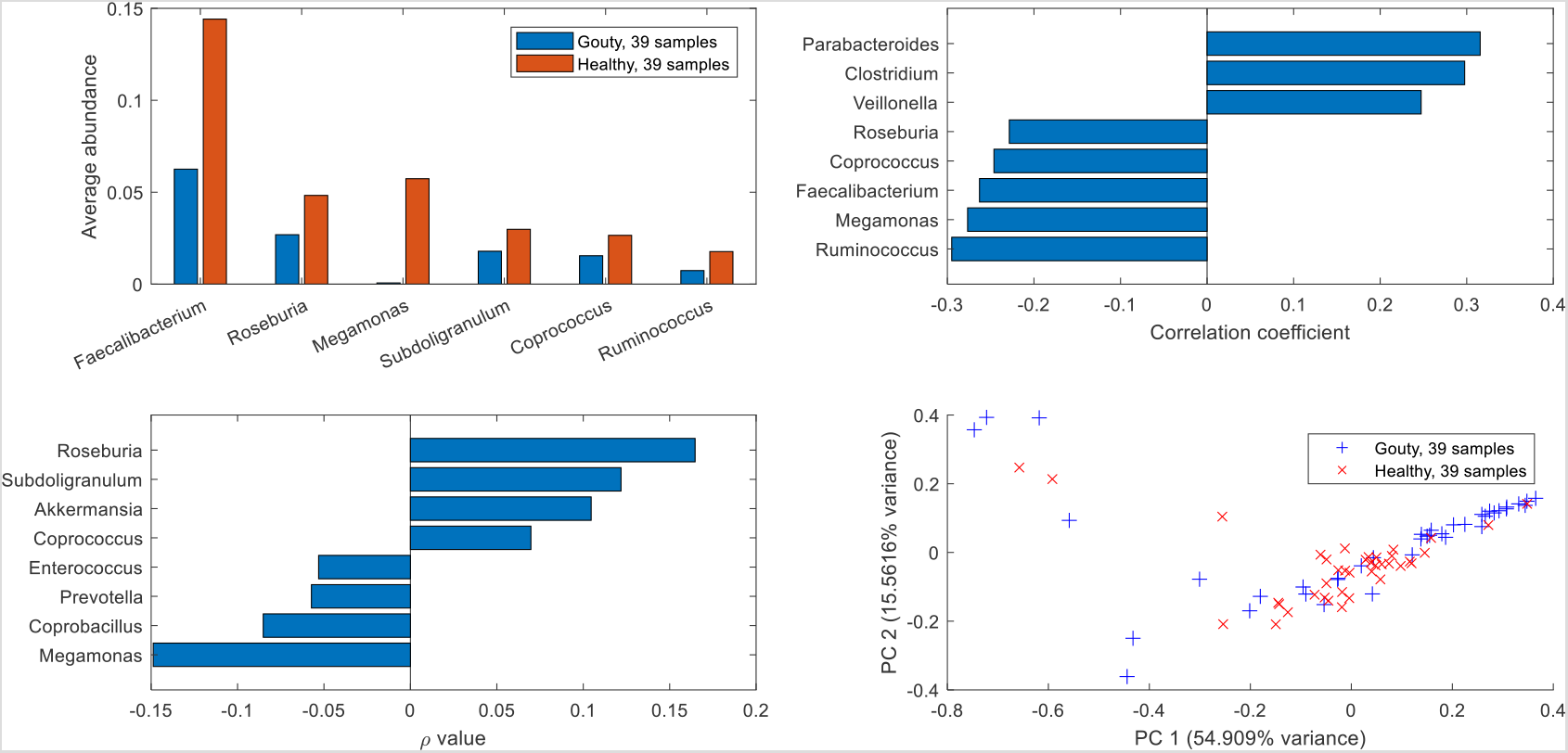
Analysis of normalized 16S-derived abundance data for samples in which the modeled taxa accounted for at least 90% of the unnormalized abundances. (A) Taxa which were significantly (FDR < 0.05) more abundant in one sample type versus the other sample type among the 25 most abundant taxa across the 78 samples used. (B) Taxa most highly correlated with the blood uric acid concentration. (C) Taxa with the highest proportionality to *Faecalibacterium*. (D) Principal component plot of the16S-derived abundance data showing gouty and healthy patient samples.

**Figure S2.**
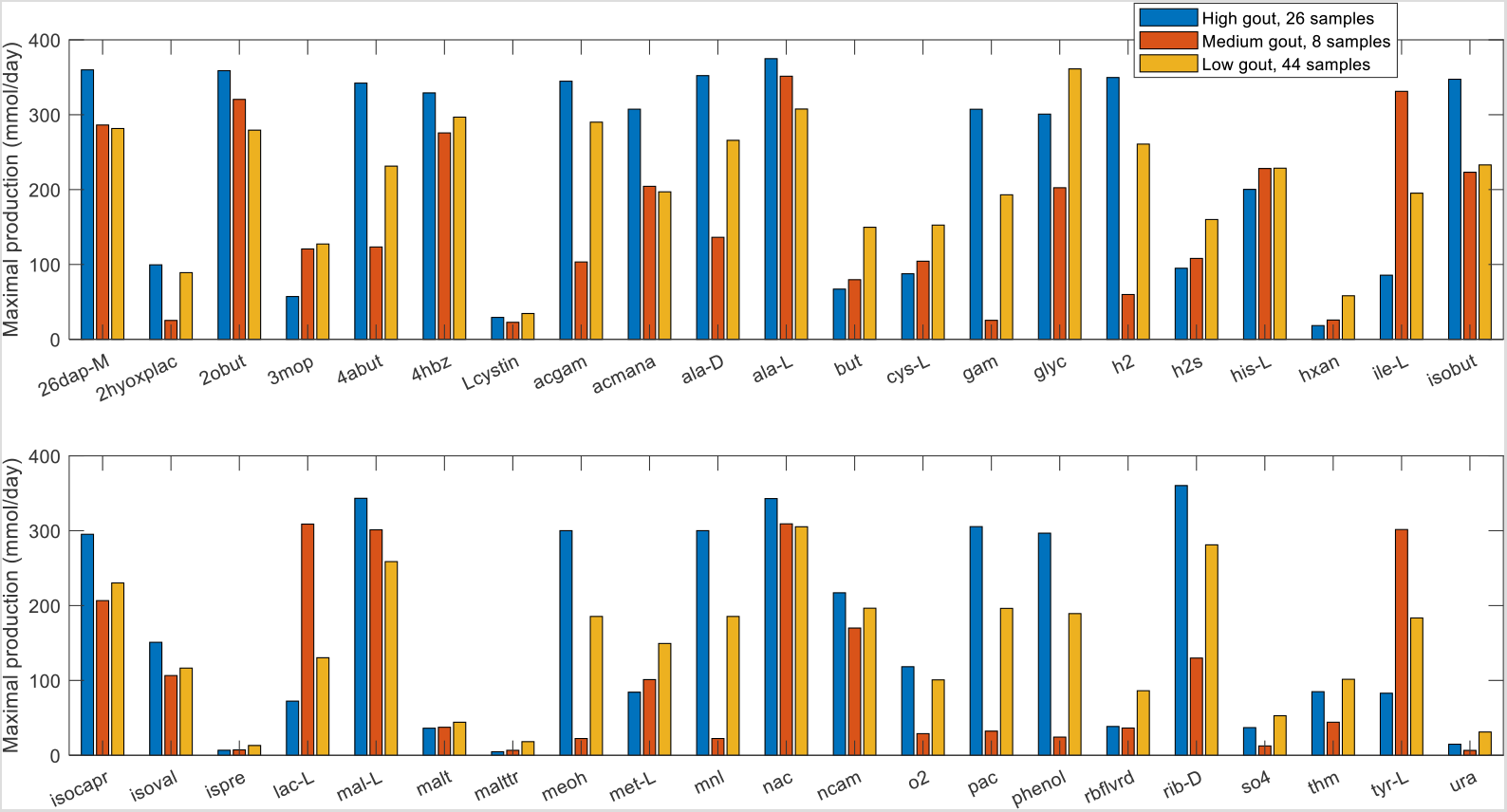
Differentially produced metabolites between the high and low gout clusters with an average EU diet. Significant differences in maximal metabolite production rates were determined by applying the Wilcoxon rank sum test (FDR < 0.05) to each metabolite across all samples in the two clusters. In addition to being statistically different, each metabolite shown had an average production rate > 10 mmol/day in at least one cluster and average production rates that differed between the clusters by at least 10%. Metabolite abbreviations are taken from the VMH database (www.vmh.life). Full metabolite names, their associated metabolic pathways and numeric values for their average production rates in each cluster are given in Table S6.

**Figure S3.**
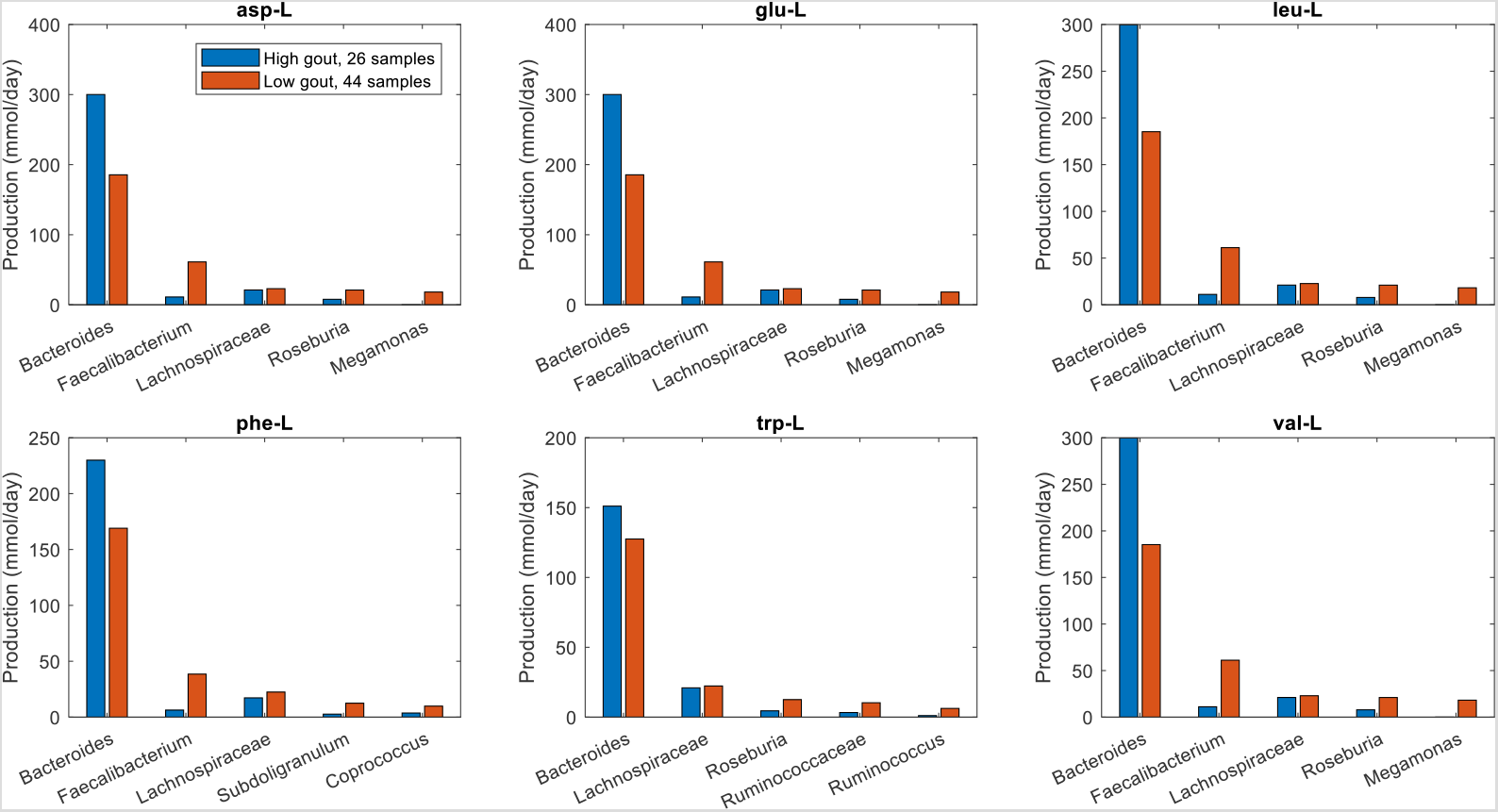
Individual taxa contributions to maximal synthesis of selected non-differentially produced amino acids in the high and low gout clusters with an average EU diet. The amino acids shown from top left to bottom right are L-aspartate, L-glutamate, L-leucine, L-phenylalanine, L-tryptophan and L-valine. For each amino acid, the top five taxa are shown in the order of their total production across the two clusters.

**Figure S4.**
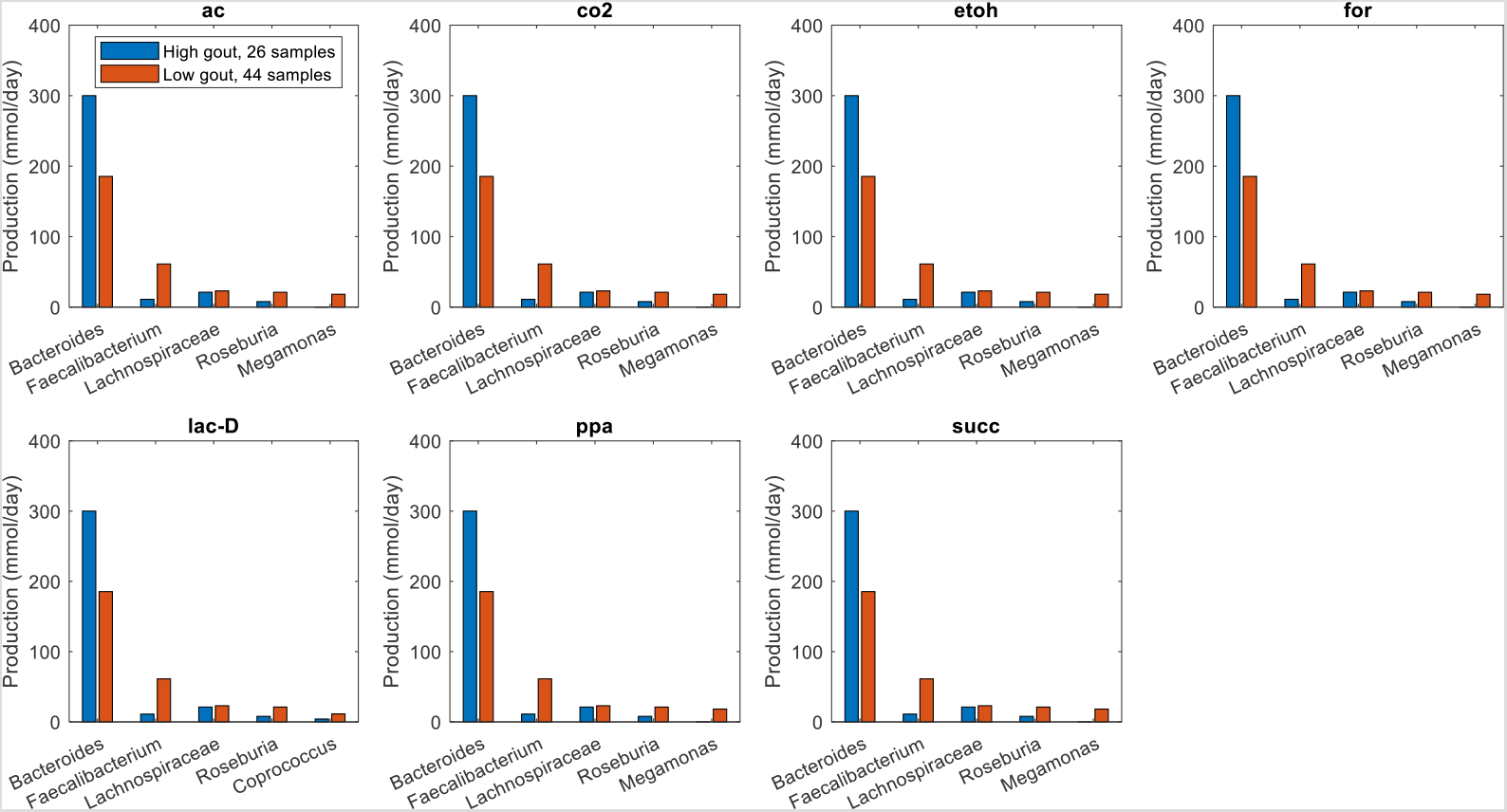
Individual taxa contributions to maximal synthesis of selected non-differentially produced fermentation byproducts in the high and low gout clusters with an average EU diet. The byproducts shown from top left to bottom right are acetate, carbon dioxide, ethanol, formate, D-lactate, propionate and succinate. For each byproduct, the top five taxa are shown in the order of their total production across the two clusters.

**Figure S5.**
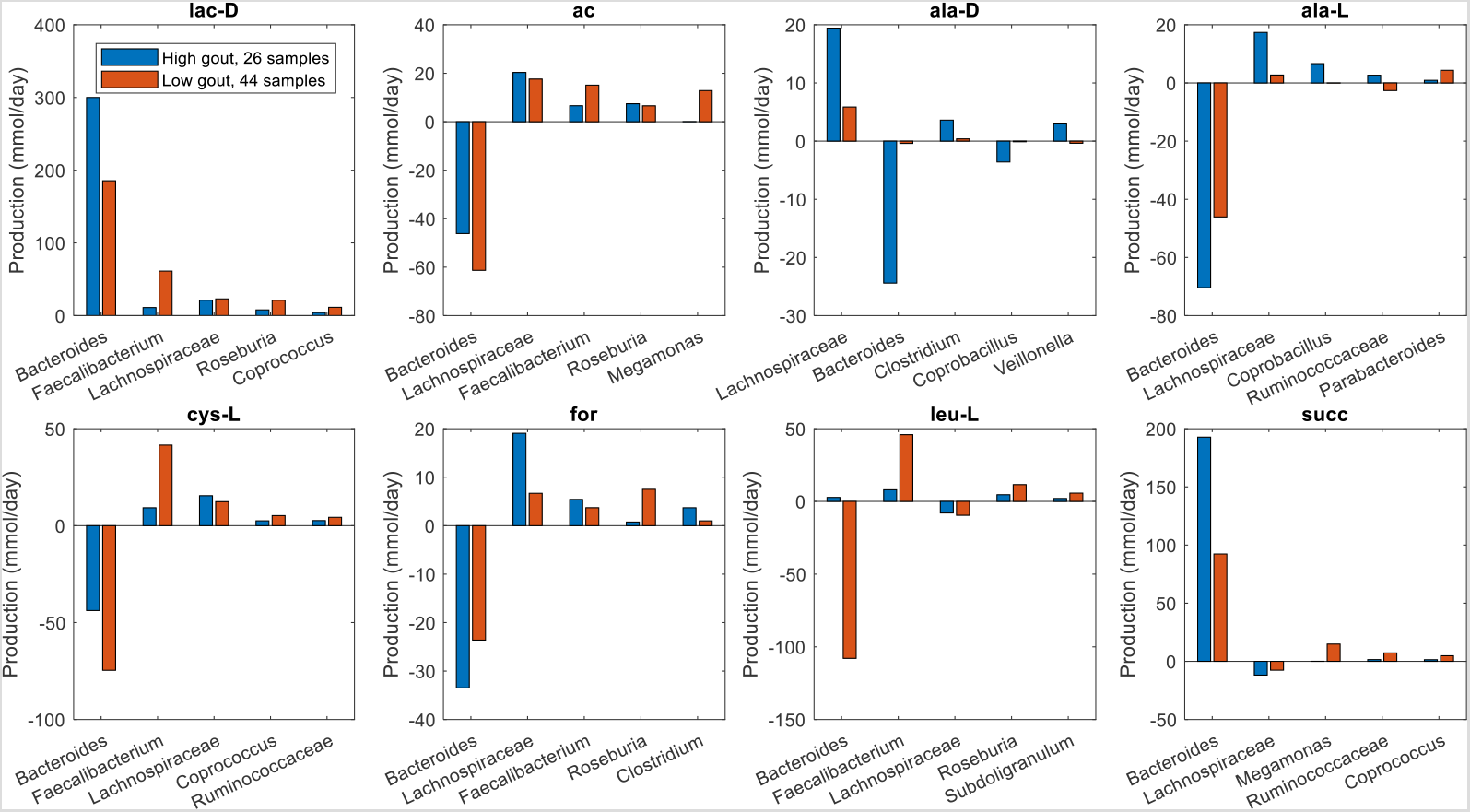
Individual taxa synthesis and uptake of crossfed metabolites for maximal D-lactate production from an average EU diet. The metabolites shown from top left to bottom right are D-lactate, acetate, D-alanine, L-alanine, L-cysteine, formate, L-leucine and succinate. Each crossfed metabolite shown had at least one taxa which satisfied minimal bounds on the metabolite secretion and uptake rates. For each metabolite, the top five taxa were ordered by the sum of the absolute values of their uptake and secretion rates across the two clusters.

**Figure S6.**
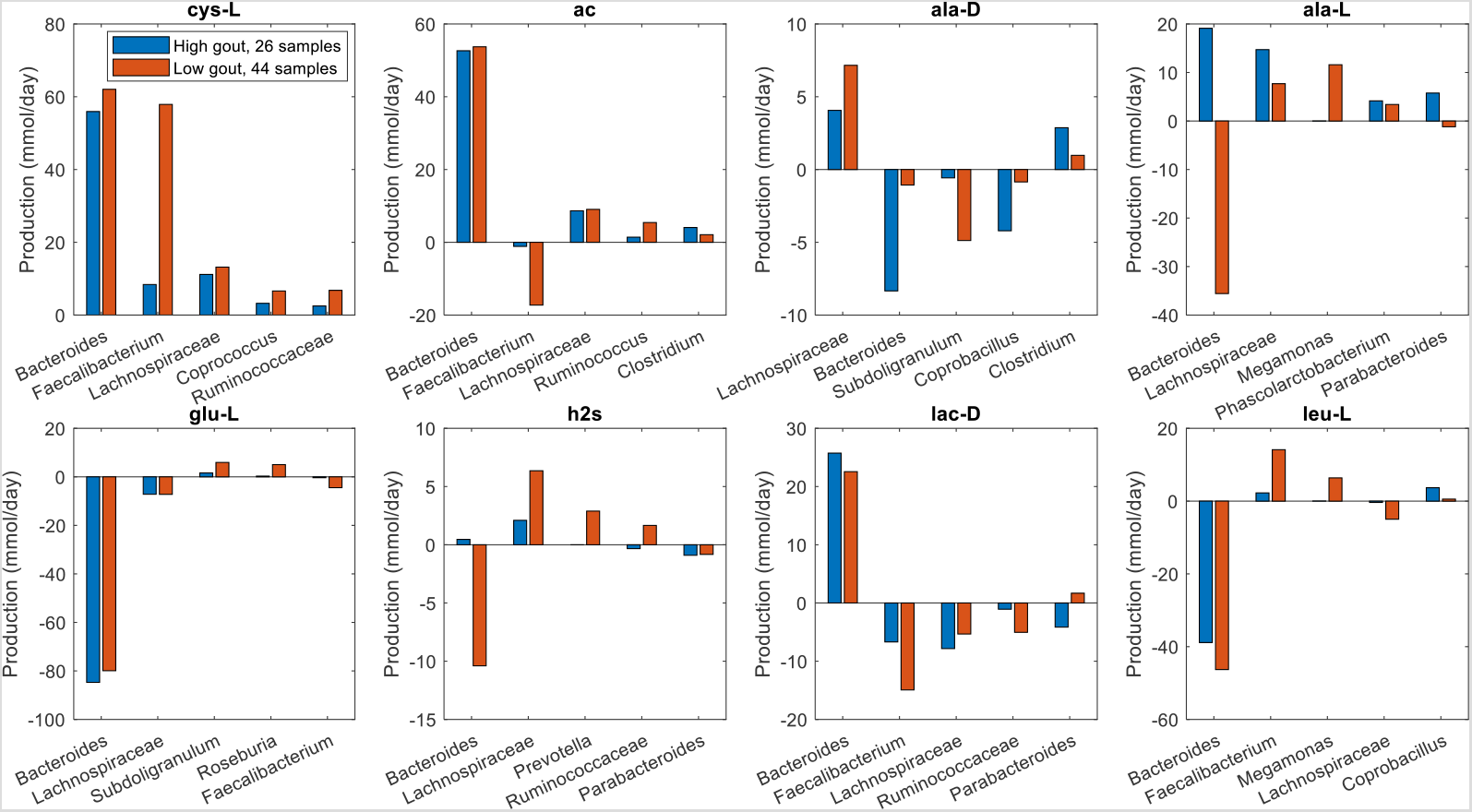
Individual taxa synthesis and uptake of crossfed metabolites for maximal L-cysteine production from an average EU diet. The metabolites shown from top left to bottom right are L-cysteine, acetate, D-alanine, L-alanine, L-glutamate, hydrogen sulfide and L-leucine. Each crossfed metabolite shown had at least one taxa which satisfied minimal bounds on the metabolite secretion and uptake rates. For each metabolite, the top five taxa were ordered by the sum of the absolute values of their uptake and secretion rates across the two clusters.

**Figure S7.**
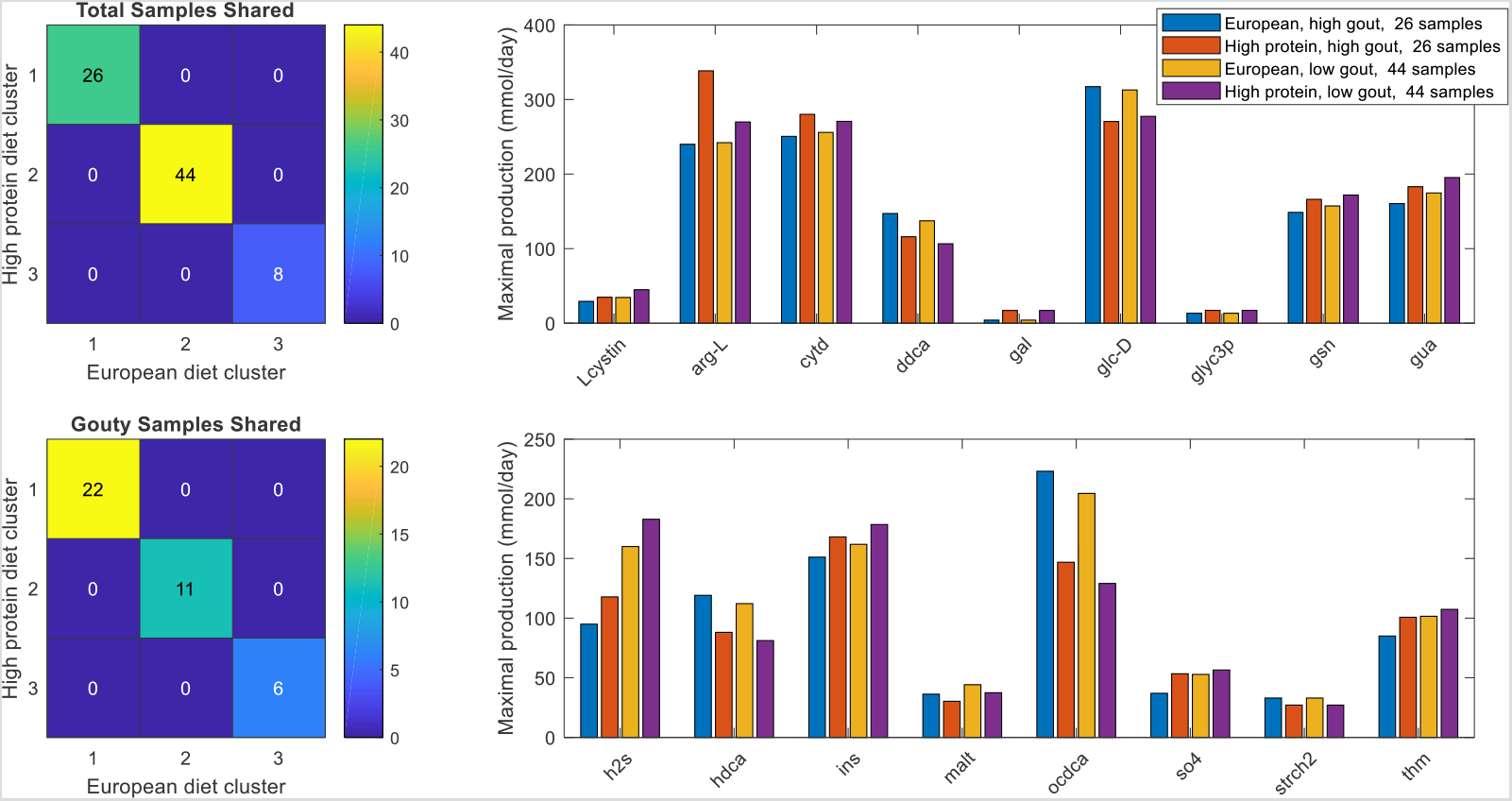
Sample clustering and differentially produced metabolites between average EU and high protein diets. (A) Total samples shared between the three clusters obtained with the average EU diet and the three clusters obtained the high protein diet. (B) Gouty samples shared between the three clusters obtained with the average EU diet and the three clusters obtained the high protein diet. (C) Significant differences in maximal metabolite production rates were determined by applying the Wilcoxon rank sum test (FDR < 0.05) to each metabolite across all samples in the two high gout clusters and the two low gout clusters. In addition to being statistically different, each metabolite shown had an average production rate > 10 mmol/day in at least one of the compared clusters and average production rates that differed between the compared clusters by at least 10%. Metabolite abbreviations are taken from the VMH database (www.vmh.life).

**Figure S8.**
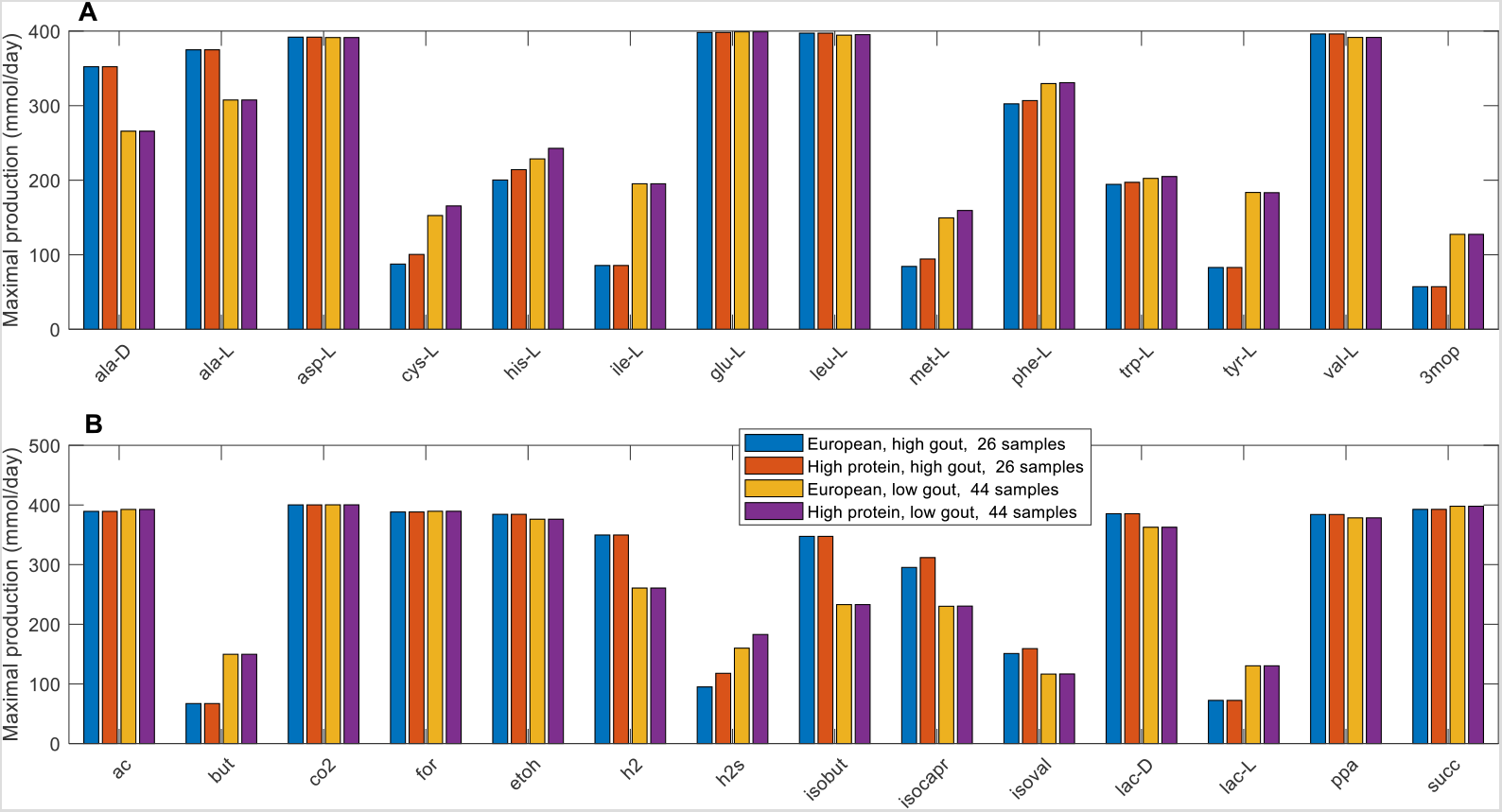
Maximal amino acid and fermentation byproduct synthesis capabilities in the high and low gout clusters from average EU and high protein diets. (A) Classes of amino acids sharing common metabolic pathways. (B) Common metabolic byproducts of carbohydrate fermentation and amino acid catabolism. Metabolite abbreviations are taken from the VMH database (www.vmh.life).

**Figure S9.**
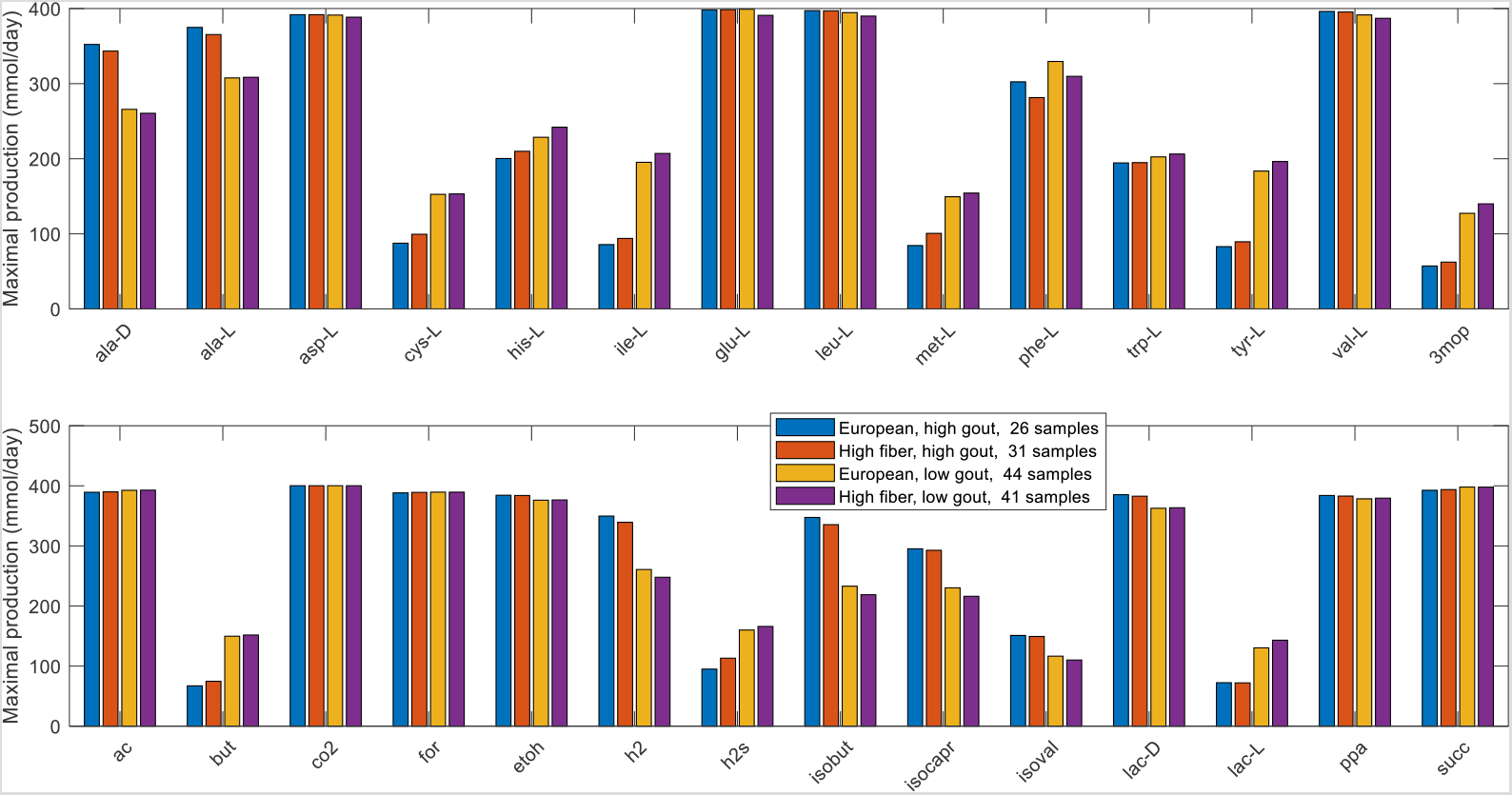
Maximal amino acid and fermentation byproduct synthesis capabilities in the high and low gout clusters from average EU and high fiber diets. (A) Classes of amino acids sharing common metabolic pathways. (B) Common metabolic byproducts of carbohydrate fermentation and amino acid catabolism. Metabolite abbreviations are taken from the VMH database (www.vmh.life).

**Figure S10.**
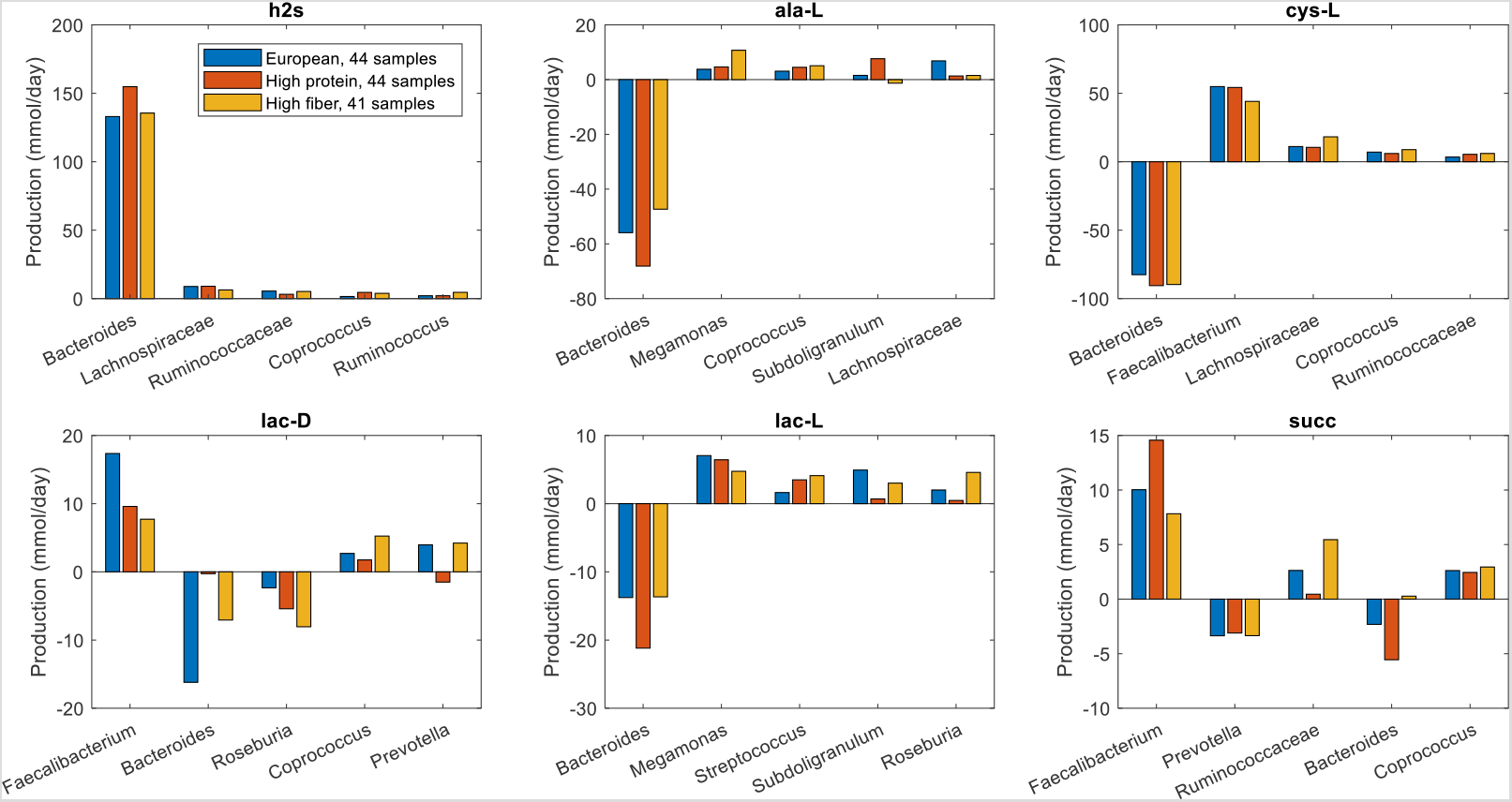
Individual taxa synthesis and uptake of crossfed metabolites for maximal H_2_S production from low gout clusters generated from average European, high protein and high fiber diets. The metabolites shown from top left to bottom right are hydrogen sulfide, L-alanine, L-cysteine, D-lactate, L-lactate and succinate. Each crossfed metabolite shown had at least one taxa which satisfied minimal bounds on the metabolite secretion and uptake rates for at least one diet. For each metabolite, the top five taxa are ordered by the sum of the absolute values of their uptake and secretion rates across the three diets.

